# Interplay between patient and centre factors in home therapy uptake: A sequences-of-regressions analysis using linked UK Renal Registry to centre survey data

**DOI:** 10.1101/2025.07.19.25331842

**Authors:** Jessica Potts, Camille M Pearse, Mark Lambie, James Fotheringham, Harry Hill, David Coyle, Sarah Damery, Kerry Allen, Iestyn Williams, Simon J Davies, Ivonne Solis-Trapala

## Abstract

**Rationale & Objective:** Disparities in home dialysis therapy (HT) use may stem from the interplay between dialysis centre services and patient characteristics. We analysed how these factors directly and indirectly affect HT uptake in England.

**Study design:** Linked UK Renal Registry (UKRR) cohort to a national survey of renal centres informed by ethnographic observation.

**Setting & Participants:** Adults who initiated kidney replacement therapy (KRT) between 2015 and 2019 at 51 English renal centres, totalling 32,400 individuals identified through the UKRR, with centre practices captured from a 2022 national survey.

**Exposures or predictors:** Patient- and centre-level factors

**Outcomes:** Use of HT (home haemodialysis or peritoneal dialysis) within one year of starting KRT.

**Analytical Approach:** Sequences of regressions, an extension of path analysis, were used to examine the direct and indirect associations between patient- and centre-level factors and the probability of HT uptake.

**Results:** Direct associations revealed that both centre- and patient-factors significantly influenced the probability of HT uptake. Patients at centres conducting quality improvement projects, (OR [95% CI]) 1.94, [1.36-2.76]), offering assisted PD (1.89, [1.39-2.57]), fostering staff research engagement (1.35, [1.03-1.77]) or hosting HT roadshows (1.22, [1.05-1.41]) had higher odds of HT uptake. Centres with staff capacity stress had lower uptake (0.60, [0.45-0.81]). Patients on transplant lists at KRT start (2.55, [2.35-2.77]) or living further from a treatment centre (1.10, [1.08-1.12] per 10km) had higher odds of HT uptake. Patients from more deprived areas or minority ethnic groups had lower HT uptake. However, an indirect association was observed through centre practices, as certain centres serving ethnically diverse populations implemented practices that directly increased HT uptake, potentially mitigating disparities.

**Limitations:** Healthcare professional-reported and aggregated survey data

**Conclusions:** This study identified modifiable centre-level factors that could improve equity in HT access and uptake by mitigating ethnic and area-level disparities in diverse populations.

## Introduction

Home dialysis therapy (HT), including peritoneal dialysis (PD) and home haemodialysis (HHD), are important for life participation, optimising survival and reducing healthcare costs, particularly as the prevalence of kidney failure continues to increase worldwide.^1^

The critical importance of shifting healthcare delivery models towards community based care has been emphasised.^2,3^ This shift is particularly relevant for managing long-term conditions such as kidney failure. Expanding HT use aligns with recommendations to support patient-centred care, enhance accessibility, and ultimately lead to better outcomes.^4^

Despite longstanding national policies recommendations favouring HT ^5,6^ and many attempts to increase uptake in high-income countries, uptake remains low.^7–11^ In England, there is significant variation in the use of HT between renal centres ^12,13^, with HT uptake particularly low in people from minority ethnic groups and deprived communities. Previous studies examining the associations between patient demographics and HT uptake in renal centres have had variable success in explaining these inequalities in service provision.^13–17^

Inter-CEPt is a sequential mixed-methods study that seeks to identify modifiable centre-level factors associated with HT uptake in England with a view to developing interventions aimed at increasing usage and reducing inequalities.^18^ We conducted an ethnographic study^19^ and used the findings to develop a national survey of centre-level characteristics, including organisational culture, practices and service organisation.^20^

In this paper, we examine the interplay between centre- and patient-level factors in shaping HT uptake. Using sequences of regressions,^21–24^ an extension of path analysis, we assess both the direct effects of patient- and centre-level factors on HT uptake, and the indirect effects, where patient factors are associated with HT uptake through differences in centre practices. By linking UK Renal Registry (UKRR) patient-level data to centre survey data, we provide novel insights into how institutional practices and patient characteristics together influence access to HT. These insights can guide quality improvement and inform policies to reduce disparities in HT uptake.

## Methods

### National survey of renal centres in England

The survey was conducted across all 51 English renal centres between June and September 2022.^20^ It was developed by reviewing existing literature and incorporating qualitative and observational data from an ethnographic study to understand factors influencing HT uptake at specific centres. It comprised 78 (dichotomous or Likert-type scale) questions across 12 sections related to centre practice patterns and the organisation of the home dialysis services. Staff involved in providing home dialysis services, including centre managers, clinical leads/directors, PD and HHD consultants and nurses, and Advanced Kidney Care clinic staff were invited to participate. It could be completed by multiple staff members per centre. The detailed steps of data aggregation, transformation, and survey question selection for analysis can be found in the Supplementary Material.

### UK Renal Registry data and data linkage

Our study population included patients (>18 years) starting KRT between 1^st^ January 2015 and 31^st^ December 2019 in England, identified through the UKRR. We used UKRR data, including demographics (age, sex, ethnicity, and neighbourhood deprivation) and clinical characteristics (diabetes as primary renal diagnosis, transplant waitlist status, distance to nearest renal centre) at KRT initiation, and treatment timelines.

Patient ethnicity was categorised in five groups based on the 2021 census: Asian, Black, Mixed, White and Other. Asian patients included those identifying as Bangladeshi, Pakistani, Indian, Chinese or “Other Asian”. Black patients included African, Caribbean and “Other Black”. Mixed patients were White and Asian, White and Black or “Other mixed”. White patients were British, Irish or “Other White”. Those identifying as Arab or “Any other ethnicity” were categorised as “Other” ^25^. Neighbourhood deprivation was measured using the Index of Multiple Deprivation (IMD)^26^ quintile score with higher quintiles indicating more deprivation.

The centre-level survey dataset was linked to the patient-level UKRR dataset by matching each patient record with their respective centre information.

### Primary outcome measure

The primary outcome was whether a patient used HT (PD or HHD) within one year of starting KRT. A patient was considered to have received HT if they used either HHD or PD at any time within the first year, for any duration. Since HT may require set-up time and training, particularly for HHD or late presenters, the period was extended to 12 months to ensure these patients were included.

### Hypothesised sequence of centre- and patient-level factors associated with HT uptake

Figure 1 shows our hypothesised sequence of interactions between centre- and patient-level factors that potentially influence HT uptake ^18^. Specifically, we hypothesised that centre-level factors, such as a centre’s approach to HT and availability of resources, may influence the patient’s access to HT. Additionally, patient-level factors such as age, sex, residential distance from the dialysis unit, ethnic group and area-level deprivation may affect the patient’s probability of having HT. The decision to start HT may be influenced by a combination of these patient characteristics and the support provided by the dialysis centre. For example, a patient from a neighbourhood with high levels of deprivation may be less likely to choose HT, but if the centre offers specific support, this could make HT more accessible. Thus, we postulated that the uptake of HT was shaped by the interplay between centre- and patient-level factors.

**Figure 1:**
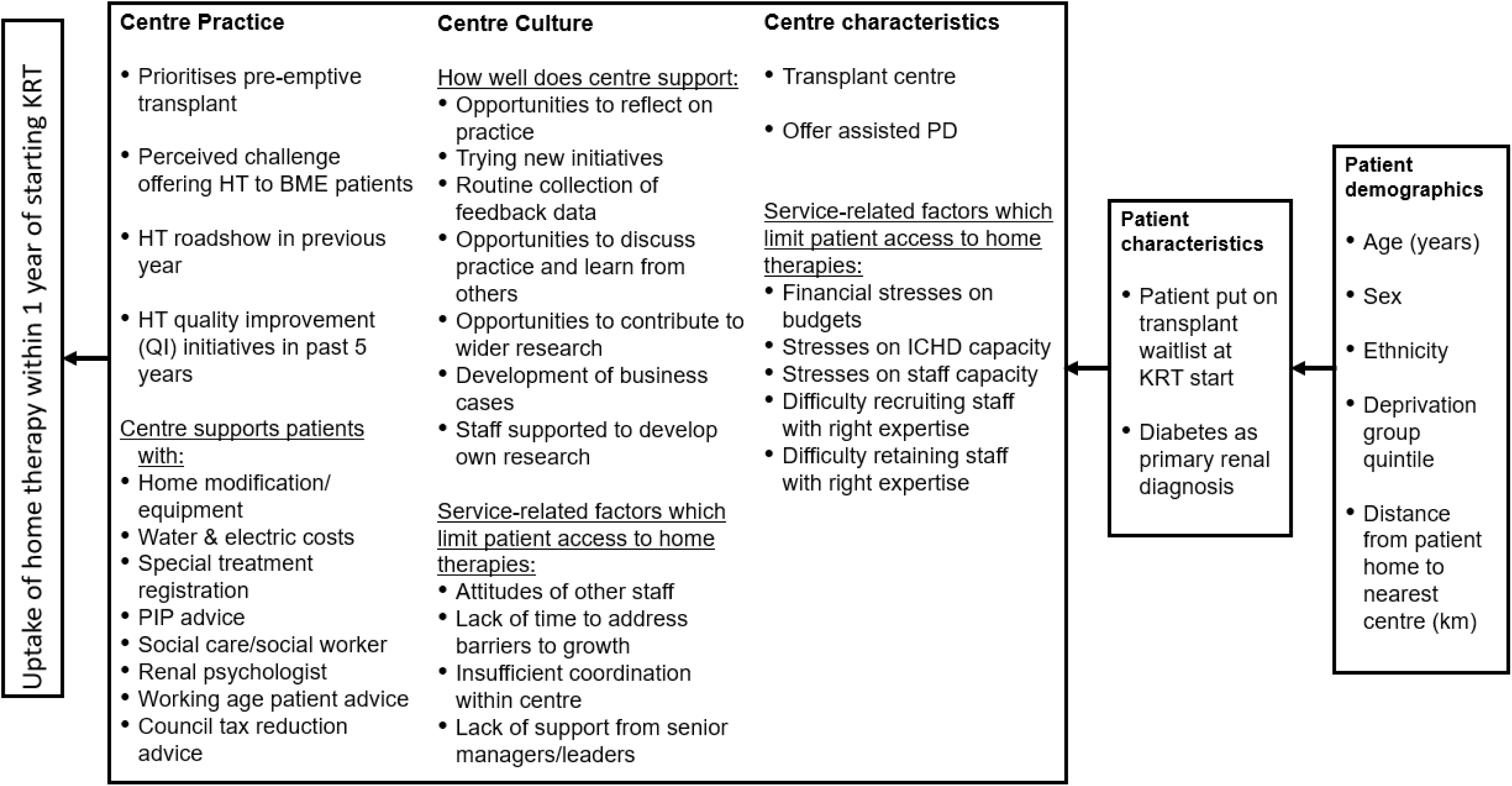
Proposed sequence of centre- and patient-level factors in their association with HT uptake within one year of starting KRT

### Statistical analysis

We used sequences of regressions^21–24^ (SoR), a graphical model that builds on path analysis, to examine the direct and indirect associations between centre- and patient-level factors and the probability of HT uptake. The model was fitted through an ordered series of regression models starting with HT uptake, the primary outcome, and working from left to right (Figure 1). HT uptake was the response variable to all the factors located within the boxes on the right-hand side and was modelled using a mixed-effects logistic regression model with a random intercept to account for centre clustering. The second box contains multiple centre-level factors which were modelled as response variables to patient characteristics and demographics, located within the third and fourth boxes using logistic regression models.

Finally, the patient characteristic variables were modelled as response variables to the patient demographics using mixed-effects logistic regression models. The best-fitting regression model for each outcome in the SoR analyses was selected by comparing nested models with different combinations of explanatory factors. The SoR are described using a regression graph in which two variables located in different boxes were linked by an arrow line emerging from a selected explanatory variable and pointing to a response variable if they are directly associated (i.e. association not explained by any of the intermediary factors). A sequence of connected arrow lines between two variables represents an indirect association (i.e. partially explained by intermediary factors). The strength of the associations depicted by the arrows in the graph were quantified using odds ratios (exponential of partial regression coefficients). SoR also allow for the exploration of residual pairwise associations of multiple factors (centre factors or patient characteristics) after accounting for their combined explanatory variables. We applied this approach to explore residual associations among centre-level factors linked with HT uptake. Further details on model interpretation, estimation, assumptions, missing data, goodness of fit, and diagnostic checks are provided in the Supplementary Material.

The level of statistical significance was set at 0.05. The analyses were carried out using STATA version 18^27^, and the statistical software R^28^.

### Patient and public involvement summary

Patients, and family members/carers, with lived experience of dialysis for kidney failure were involved at every stage of the Inter-CEPt study including design, application for grant funding, study management, interpretation and dissemination of findings. A dedicated patient advisory group, supported by the Keele University Patient and Public Involvement team, representative of diverse ethnicities, geographies and backgrounds met seven times over the course of the project and co-produced the final public-facing report of the research.

### Ethical approval and informed consent

Ethical approval for this study was granted by the UK Health Research Authority (Ref: 20-WA-0249). Centre survey participants provided informed consent via an embedded form at the start of the survey. Pseudonymised patient data were provided by the UKRR under study approval (Ref: DSA93). The UKRR holds Section 251 approval under the NHS Act 2006 to process and share confidential patient data for research, with ethics approval from the Research Ethics Committee (Ref: 16/NE/0042).

## Results

### Survey data

A total of 180 responses were received from 50 of 51 kidney centres ^20^, with 1-10 responses per centre (average 3.5). We selected 43 questions relevant to both PD and HHD, deriving 98 factors from them since each question addressed multiple factors. After identifying potentially modifiable centre factors that could affect HT uptake and excluding factors with more than 10% missing data, 31 factors were included in the analysis (Figure S1, Table 2). Table S3 shows the patterns of missingness across centres. Of the 50 centres, 38 (76%) had complete responses for all factors.

#### UKRR data

Of the 32,400 incident KRT patients between 2015 and 2019, 72% started on in-centre HD (ICHD), (n=23,242), 20% on HT (n=6,522) and 8% had a pre-emptive kidney transplant (n=2,636). Within the first calendar year of starting KRT, 8,147 (25%) patients had received HT. Table 1 shows a summary of the patient characteristics stratified by initial KRT modality. Patients starting on HT were younger than those starting on ICHD. A greater proportion of patients from areas of least deprivation received HT or transplant as their initial modality. The proportion of patients who received HT and were waitlisted for transplant at start of KRT was 3-fold that of those who started on ICHD. Tables S1 and S2 show the incident KRT patients by renal centre and year, and patient demographics over the study period, respectively. The proportion of patients receiving HT within one year varies between centres and there is no apparent pattern across centre sizes, defined as the proportion of the incident cohort (range: 0.5% to 5.7%). Larger centres tend to have more ethnically diverse populations. Missing data across the variables used in the SoR analyses ranged from 0% to 8% (median: 0%, interquartile range: 0% to 4%).

**Table 1:** Patient characteristics for incident KRT patients between 1^st^ January 2015 and 31^st^ December 2019 (n=32,400) collected from the UKRR.

|  | ICHD<br>(n=23,242) (72%) |  | Home Therapy<br>(n=6,522) (20%) |  | Pre-Emptive<br>Transplant<br>(n=2,636) (8%) |  |
| --- | --- | --- | --- | --- | --- | --- |
|  | n | % | n | % | n | % |
| Age (years), median [IQR] | 66 [54,76] |  | 61 [48,72] |  | 51 [41,61] |  |
| Distance (km) from closest centre, median [IQR] | 5.8 [3.3,11.5] |  | 6.6 [3.7,12.8] |  | 7.2 [4.2,13.5] |  |
| Sex |  |  |  |  |  |  |
| Male | 14,960 | 64.4% | 4,138 | 63.5% | 1,521 | 57.7% |
| Female | 8,282 | 35.6% | 2,384 | 36.5% | 1,115 | 42.3% |
| IMD Quintile |  |  |  |  |  |  |
| (Least deprived) 1 | 3,112 | 13.4% | 1,108 | 17.0% | 595 | 22.6% |
| 2 | 3,746 | 16.1% | 1,234 | 18.9% | 563 | 21.4% |
| 3 | 4,549 | 19.6% | 1,287 | 19.7% | 551 | 20.9% |
| 4 | 5,558 | 23.9% | 1,431 | 21.9% | 468 | 17.8% |
| (Most deprived) 5 | 6,261 | 26.9% | 1,460 | 22.4% | 434 | 16.5% |
| Missing | 16 | 0.1% | 2 | 0.0% | 25 | 1.0% |
| Ethnicity |  |  |  |  |  |  |
| Asian | 3,227 | 13.9% | 906 | 13.9% | 304 | 11.5% |
| Black | 1,913 | 8.2% | 508 | 7.8% | 65 | 2.5% |
| Mixed | 363 | 1.6% | 122 | 1.9% | 59 | 2.2% |
| Other | 390 | 1.7% | 118 | 1.8% | 25 | 1.0% |
| White | 16,269 | 70.0% | 4,670 | 71.6% | 2,106 | 79.9% |
| Missing | 1,080 | 4.7% | 198 | 3.0% | 77 | 2.9% |
| Waitlisted for transplant at start of KRT |  |  |  |  |  |  |
| No | 21,371 | 92.0% | 4,911 | 75.3% | N/A | N/A |
| Yes | 1,871 | 8.1% | 1,611 | 24.7% | N/A | N/A |
| Missing | 0 | 0.0% | 0 | 0.0% | N/A | N/A |
| Diabetes as primary diagnosis |  |  |  |  |  |  |
| No | 14,954 | 64.3% | 4,507 | 69.1% | 2,115 | 80.2% |
| Yes | 6,603 | 28.4% | 1,750 | 26.8% | 340 | 12.9% |
| Missing | 1,685 | 7.3% | 265 | 4.1% | 181 | 6.9% |

IMD: Index of multiple deprivation; KRT: Kidney Replacement Therapy; ICHD: In-centre Haemodialysis

**Table 2:** Table of the number of renal unit responses in English renal survey for variables selected for analysis.

| Survey Question | No/<br>Disagree | % | Yes/<br>Agree | % | Missing | % |
| --- | --- | --- | --- | --- | --- | --- |
| Was there a HT roadshow* in previous year? | 48 | 96% | 2 | 4% | 0 | 0% |
| Have there been QI initiatives in past 5 years? | 6 | 12% | 42 | 84% | 2 | 4% |
| Is the centre a transplant centre? | 32 | 64% | 18 | 36% | 0 | 0% |
| Does the centre offer assisted PD? | 8 | 16% | 42 | 84% | 0 | 0% |
| Is it challenging to offer HT to BME patients? | 40 | 80% | 6 | 12% | 4 | 8% |
| Does the centre offer patients support with? |  |  |  |  |  |  |
| Home and equipment purchase support | 9 | 18% | 37 | 74% | 4 | 8% |
| Water and electricity cost support | 3 | 6% | 43 | 86% | 4 | 8% |
| Special treatment registration support | 3 | 6% | 43 | 86% | 4 | 8% |
| Offer PIP advice | 4 | 8% | 42 | 84% | 4 | 8% |
| Social worker/care within centre | 14 | 28% | 32 | 64% | 4 | 8% |
| Renal psychologist within centre | 14 | 28% | 33 | 66% | 3 | 6% |
| Advice for working age patients | 3 | 6% | 42 | 84% | 5 | 10% |
| Advice on council tax reduction | 3 | 6% | 42 | 84% | 5 | 10% |
| Do the following service-related factors limit patient access to HT? |  |  |  |  |  |  |
| Financial stress on centre budgets | 32 | 64% | 18 | 36% | 0 | 0% |
| Stresses on ICHD capacity | 22 | 44% | 27 | 54% | 1 | 2% |
| Stresses on staff capacity | 10 | 20% | 40 | 80% | 0 | 0% |
| Difficulty recruiting staff with correct expertise | 10 | 20% | 40 | 80% | 0 | 0% |
| Difficulty retaining staff with correct expertise | 16 | 32% | 34 | 68% | 0 | 0% |
| Attitudes of other staff in centre | 28 | 56% | 22 | 44% | 0 | 0% |
| Lack of time to address barriers to growth | 20 | 40% | 30 | 60% | 0 | 0% |
| Insufficient coordination within renal centre | 36 | 72% | 13 | 26% | 1 | 2% |
| Lack of support from senior managers/leaders | 35 | 70% | 15 | 30% | 0 | 0% |
| Does the centre support the following? |  |  |  |  |  |  |
| Opportunities to reflect on practice | 5 | 10% | 44 | 88% | 1 | 2% |
| Encouraging new initiatives | 5 | 10% | 44 | 88% | 1 | 2% |
| Routine collection of feedback data | 16 | 32% | 34 | 68% | 0 | 0% |
| Discuss practice and learn from others | 9 | 18% | 41 | 82% | 0 | 0% |
| Opportunities to contribute to wider research | 13 | 26% | 37 | 74% | 0 | 0% |
| Support for developing business cases | 14 | 28% | 35 | 70% | 1 | 2% |
| Support staff to develop own research | 23 | 46% | 27 | 54% | 0 | 0% |

\*An HT roadshow is an initiative whereby a dialysis centre is visited by a team including patients, family members, clinicians and industry to promote HT; HT: Home therapy; QI: Quality improvement; BME: Black and minority ethnic; ICHD: In-centre Haemodialysis; PD: Peritoneal dialysis, PIP: Personal independence payment (PIP provides help with extra living costs for people who have difficulty doing everyday tasks due to having a long term health condition or disability)

#### Direct associations of patient- and centre-level characteristics with home therapy uptake

Table 3 presents the estimated direct associations of centre- and patient-level factors with HT uptake from the model of best fit. Figure 2 shows a subgraph displaying these associations alongside indirect associations.

**Figure 2:**
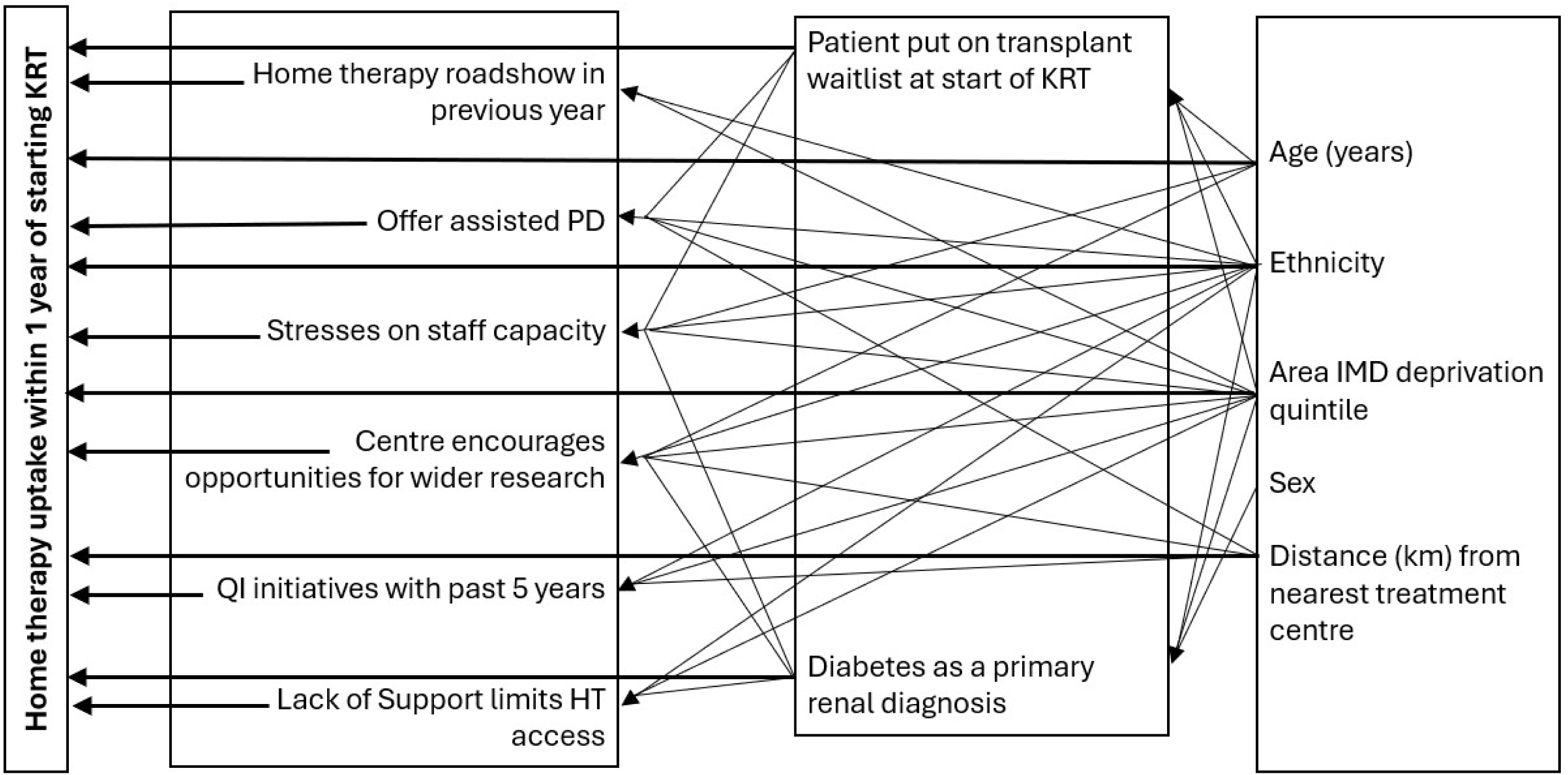
Regression subgraph of direct and indirect associations of centre- and patient-level factors with HT uptake Two variables in separate boxes were connected by an arrow line, emerging from a selected explanatory variable and pointing to a response variable if they are directly associated (i.e. association not explained by any of the intermediary factors). Direct associations with HT uptake are highlighted in bold. A sequence of connected arrow lines between two variables represents an indirect association (i.e. partially explained by intermediary factors). All centre level factors showed strong pairwise associations, after controlling for their combined set of explanatory variables (lines between any two factors are not shown in the graph to maintain clarity and avoid overcrowding).

**Table 3:** Direct associations of centre- and patient-level factors with the probability of a HT uptake within one year of commencing KRT based on mixed-effects logistic regression model of best fit.

| Factor Level | Descriptor | Odds ratio (95% CI) |
| --- | --- | --- |
| Patient demographic | Age (per 10-year increase at KRT start) | 0.91 (0.90-0.92) |
|  | Ethnicity |  |
|  | White (reference) | - |
|  | Asian | 0.84 (0.77-0.92) |
|  | Black | 0.84 (0.75-0.95) |
|  | Mixed | 0.77 (0.62-0.96) |
|  | Other | 0.93 (0.75-1.16) |
|  | IMD quintile |  |
|  | (least deprived) 1 | 1.34 (1.21-1.48) |
|  | 2 | 1.17 (1.06-1.28) |
|  | (Reference) 3 | - |
|  | 4 | 0.86 (0.79-0.95) |
|  | (most deprived) 5 | 0.74 (0.68-0.81) |
|  | Living distance from centre (per 10km) | 1.10 (1.08-1.12) |
| Patient characteristics | On the transplant waiting list at start | 2.55 (2.35-2.77) |
|  | Diabetes as primary diagnosis | 0.93 (0.88-0.99) |
| Centre characteristics | Centre offers assisted PD | 1.89 (1.39-2.57) |
|  | Stresses on staff capacity limits HT access | 0.60 (0.45-0.81) |
| Centre culture | Opportunities to contribute to research | 1.35 (1.03-1.77) |
|  | Lack of support limits HT access | 1.47 (1.13-1.92) |
| Centre practices | Home dialysis related Quality Improvement (QI) initiative in the last 5 years | 1.94 (1.36-2.76) |
|  | HT Roadshow* in the last year | 1.22 (1.05-1.41) |

\*An HT roadshow is an initiative whereby a dialysis centre is visited by a team including patients, family members, clinicians and industry to promote HTs; IMD: Index of Multiple Deprivation; HT: Home therapy; KRT: Kidney replacement therapy; PD: Peritoneal dialysis; QI: Quality improvement

#### Centre-level factors

Higher odds of HT uptake were linked to renal centres that conducted quality improvement (QI) projects in the last five years (odds ratio (OR), [95% CI]) (1.94, [1.36-2.76]), offered assisted PD 1.89 [1.39-2.57], fostered research (1.35, [1.03-1.77]) or hosted home dialysis roadshows (1.22, [1.05-1.41]). Other centre-level variables that influenced HT uptake included stress on staff capacity to deliver HT (0.60, [0.45-0.81]) and a perceived lack of support from senior managers/leaders which limited opportunities for HT (1.47, [1.13-1.92]). These associations are depicted in Figure S2 through predicted HT uptake probabilities.

Additionally, we identified two potential interaction terms on the probability of HT uptake: between research opportunities and perceived lack of support which limits HT, and between research opportunities and running QI projects. However, due to data sparsity, inclusion of these interactions led to wide confidence intervals in regression estimates, indicating a high degree of uncertainty (see Supplementary Material).

#### Patient-level characteristics

The odds of HT uptake were 2.6-fold for patients waitlisted for transplant at the start of KRT (2.55, [2.35-2.77]). For patients living further from the nearest treatment centre by 10 km, the odds of HT uptake were 10% higher (1.10, [1.08-1.12]). Patients with diabetes as the primary cause of renal disease had 7% lower odds of HT uptake (0.93, [0.88-0.99]). Additionally, with a decade difference in age at KRT initiation, the odds of HT uptake were 9% lower (0.91, [0.90-0.92]).

Patients from Asian (0.84, [0.77-0.92]), Black (0.84, [0.75-0.95]), Mixed (0.77, [0.62-0.96]) groups had lower odds of HT uptake compared to White patients. Compared to the reference 3^rd^ quintile of deprivation, patients from lower deprivation areas (quintiles 1 and 2) had higher odds of HT uptake (1.34, [1.21-1.48] and 1.17, [1.06-1.28] respectively), while those from higher deprivation areas (quintiles 4 and 5) had lower odds (0.86, [0.79-0.95], 0.74, [0.68-0.81]). Figures S3 and S4 illustrate these associations through predicted HT uptake probabilities.

#### Indirect associations of patient-level characteristics with home therapy uptake

Figure 2 and Table 4 describe how patient-level characteristics are associated with centres that adopt specific practices, indicating potential indirect effects on HT uptake.

**Table 4:**
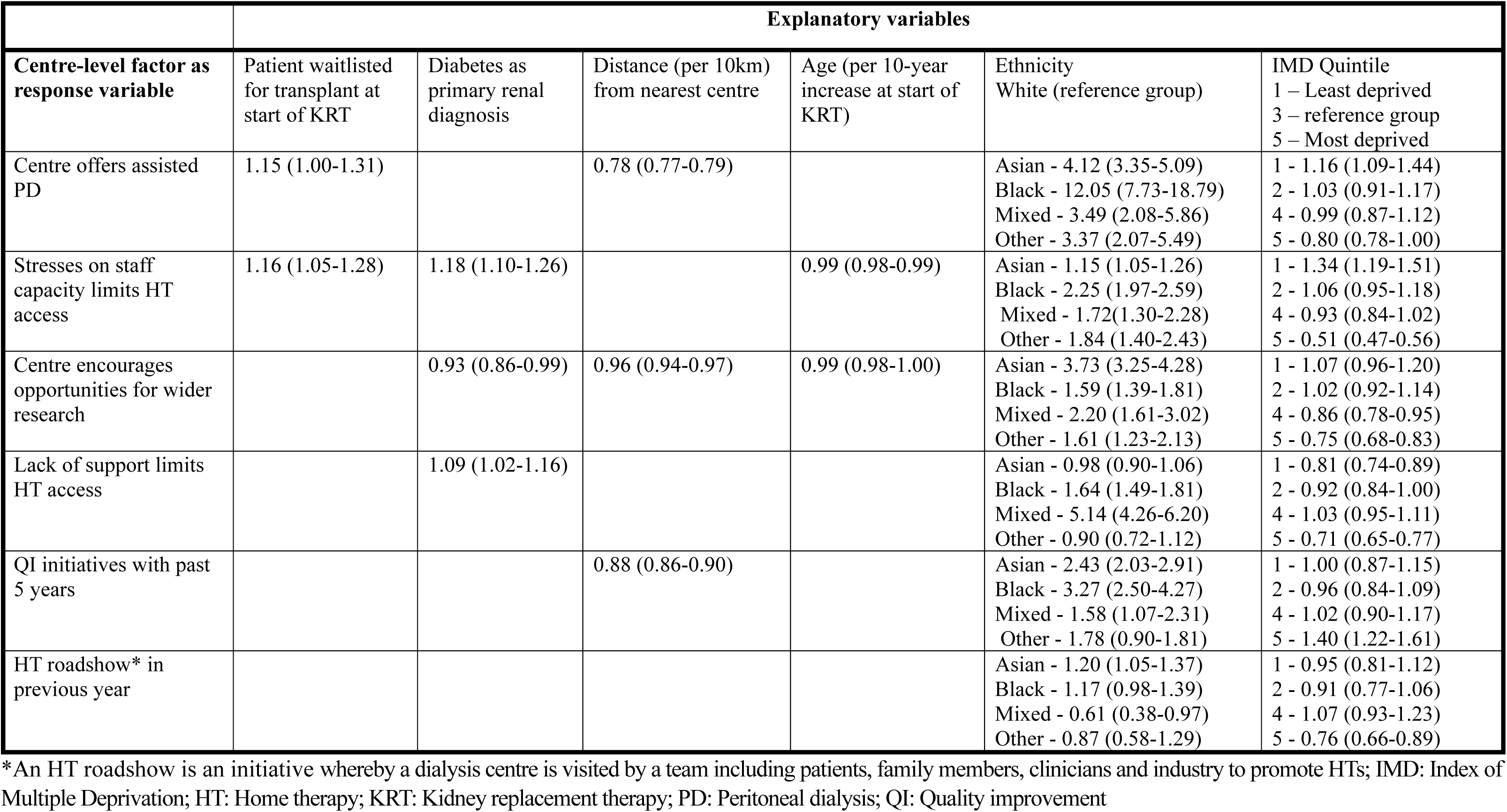
Indirect associations: Associations between patient-level characteristics and centre-level characteristics that were directly related to HT uptake based on the logistic regression models of best fit.

Centres that have implemented QI initiatives within the past five years tend to have higher proportions of patients from the most deprived areas (IMD quintile 5) compared to quintile 3 (1.40, [1.22-1.61]) and patients identifying as Asian or Black (2.43, [2.03-2.91] and 3.27, [5.50-4.27] respectively) compared to White patients. Centres that hosted an HT roadshow in the previous year similarly show a higher presence of Asian patients (1.20, [1.05-1.37]). Centres encouraging staff research opportunities are also more likely to have a diverse patient base, including higher proportion of Asian, Black, Mixed and Other ethnic groups compared to White patients. Conversely, the proportion of patients from higher deprivation areas (Quintile 4: 0.86, [0.78-0.95], Quintile 5: 0.75, [0.68,0.83]), and those with diabetes as primary diagnosis (0.93, [0.86-0.99]) are lower in these centres.

Centres offering assisted PD had higher proportion of patients who were initially waitlisted for KRT (1.15, [1.00-1.31]), from areas with the least deprivation (1.16, [1.09-1.44]) compared to those in the quintile 3 group, and belonging to Asian (4.12, [3.35-5.09]), Black (12.05, [7.73-18.79]), Mixed (3.49, [4.08-5.86]) or Other ethnic groups (3.37, [2.07-5.49]) compared to White patients. Centres experiencing stress on staff capacity had higher proportions of waitlisted transplant patients (1.16, [1.05,1.28]), those with diabetes as primary renal diagnosis (1.18, [1.10-1.26]) and patients from less deprived areas. Additionally, centres with perceived lack of staff support saw higher proportions of patients with diabetes as primary diagnosis (1.09, [0.102-1.16]) and those identified as Black (3.05, [2.66-3.52]) or Mixed ethnicity (5.14, [4.26-6.20]).

Further regression models for centre-level and patient-level factors from the SoR analysis are presented in the Supplementary Material (Tables S5-S6)

## Discussion

This study offers the most comprehensive analysis to date of the interplay between centre practices, indicators of organisational culture and patient characteristics influencing the probability of home therapy uptake. Using an innovative approach based on sequences of regressions applied to registry data linked to centre national survey data in England, it constitutes a central component of a sequential mixed-methods design that will inform a service delivery intervention designed to reduce the centre-level variation in HT ^18^.

Our findings suggest that renal centres running quality improvement projects on home dialysis, hosting home dialysis road shows, fostering staff research engagement, and offering assisted PD were directly associated with higher odds of HT uptake. Conversely, centres in which there is perceived stress on staff capacity had lower HT uptake. Known patient demographic factors were confirmed, including lower odds of being on HT amongst ethnic minority groups compared to White patients, older patients or those from higher deprivation areas according to the IMD.

While the overall trend shows that patients from minority ethnic groups generally experience lower HT uptake, our analysis reveals that certain centres serving ethnically diverse populations have adopted practices that effectively enhance HT uptake among these groups. This suggests that disparities at the individual patient level, while prevalent, can be mitigated through specific practices implemented within these centres. Collectively, our findings of both direct and indirect associations not only highlight these effective practices but also underscore their potential to inform targeted interventions designed to address inequities in HT access.

Our findings build on previous research into how centre characteristics and physician practice patterns are associated with home dialysis use.^17^ The patient factors identified by Castledine *et al.* were broadly similar, although the number and type of centre level factors was more restricted, differently defined or differently associated with HT use, in part because clinical practices have changed over time. For example, the availability of assisted PD was not investigated previously because its use was not common in 2013, but since then it has become more widespread and funded by a specific reimbursement tariff in England. This analysis provides the first national-level evidence that the use of assisted PD increases access to HT, aligning with single-centre data^29^ and more informal survey data across Europe.^30^ Equally, the ease with which a PD catheter can be inserted was previously associated with increased odds of having HT whereas we observed no such association in our survey^20^ as this measure did not get into the model because of a lack of variation between centres. Again, this may reflect the development in services in response to the expectations of commissioners of dialysis in England.

Castledine *et al.* also reported a strong association between the physician’s enthusiasm for HT and its use and indeed this observation was one of the reasons why we initiated the Inter-CEPt study with an ethnography to understand what this meant.^17^ Our finding that pro-HT leadership translates into a strong pro-HT organisational culture led to our inclusion of several aspects that typified this centre characteristic in our study ^19^. Our finding that several of these factors are associated with greater use of HT does not contradict the importance of physician enthusiasm. Instead, it reveals the effects of this enthusiasm and how it might be emulated. A recent study in Australia found that centres with fewer patients tended to have lower rates of patients on HT within six months of starting treatment.^16^ However, our survey did not identify any correlation between centre size and HT use, consistent with previous observations in the UK, possibly due to the fact that dialysis centres are relatively large.

This study was conducted within a healthcare service free at the point of care and funded through general taxation, where healthcare professionals have no obvious financial incentives affecting modality selection. The issues identified are therefore likely to be valid in similar healthcare systems, although the strength of associations may differ. Given that unwarranted variation in practices and outcomes seems to be a universal feature of healthcare, the findings may also be relevant in other healthcare systems, although the associations may be relatively weak where financial incentives have a dominant effect.

A strength of this study is that we used the UK Renal Registry cohort, which provides a representative, rich source of information about all KRT patients in England. By using a sequences of regressions analysis, we advanced previous research by not only examining the effect of centre and patient-level factors on HT uptake, but also by disentangling the complex interrelationships among these factors. This approach allowed us to separate direct from indirect associations, offering a more comprehensive understanding of the multiple influences at play in a real-world context with many contributing factors. There are, however, several limitations to our study. The analysis was partly based upon self-reported information in the survey which is subject to error as outlined in our descriptive publication.^16^ Some of the survey questions did not reach the threshold for inclusion in the sequences of regressions analysis due to missingness or lack of variation of responses. Furthermore, the study used aggregate survey scores due to multiple responses per centre, which may also reduce the influence of strong opinions. Although we identified two potential interaction terms in the regression model for HT uptake (between research opportunities for staff and perceived HT lack of support from senior leadership, and research opportunities for staff and QI initiatives in the past years), we chose to interpret a model that excluded these interactions due to the high uncertainty they introduced, as evidenced by wide confidence intervals arising from sparse data. While the main effects provide insights into the direct associations, future research should robustly evaluate these potential interaction effects. Finally, there should always be caution in inferring causal relationships from observational data. In the sequences of regressions approach, we postulated a direction of associations to explore how centre’s demographics may influence HT uptake through centre-level factors. However, causality should not be inferred, as the true direction may differ in some cases. For instance, being waitlisted at the start of KRT is linked to a transplant centre; however, causality flows from transplant centre to patient status.

Our analysis has identified several factors associated with HT uptake that are potentially modifiable, and these have been taken forward to inform the development of a service delivery intervention bundle. This includes relatively straight forward components such as recommending the use of assisted PD and HT roadshows, a process whereby a dialysis centre is visited by a team including patients, family members, clinicians and industry to promote HT modalities. More importantly it emphasises the relative importance of organisational culture over service structures or configurations. This suggests the need for interventions that tackle the factors that contribute to cultures of learning and improvement within renal centres.

## Data Availability

The data used in this study were obtained by linking centre survey data with registry data provided by the UKRR. Access to UKRR data is governed by strict data protection regulations, and the data cannot be made publicly available. To apply for access to UKRR data for research purposes visit https://www.ukkidney.org/audit-research/how-access-data/ukrr-data/apply-access-ukrr-data. The deidentified and aggregated survey data underlying the results presented in this manuscript are available by contacting the survey lead at University of Birmingham (Sarah Damery) on reasonable request from bona fide researchers with a methodologically sound proposal and the appropriate ethnical approvals. Any relevant analysis can be done on these data, which will be available for five years following manuscript publication.

## Authors’ Contributions

Research idea and study design: SJD, DC, JF, ML, IS-T; data acquisition: SD, SJD and ML oversaw the planning, design and collection of the survey data as part of the Inter-CEPt study; ML, CMP and JP facilitated access to the registry data; CMP and JP curated the data, performed the data linkage process and ensured data integrity; analysis/interpretation: JP, CMP, ML, JF, HH, DC, SD, KA, IW, SJD, IS-T; statistical analysis: JP, CMP, IS-T; supervision or mentorship: IS-T. Each author contributed important intellectual content during manuscript drafting or revision and agrees to be personally accountable for the individual’s own contributions and to ensure that questions pertaining to the accuracy or integrity of any portion of the work, even one in which the author was not directly involved, are appropriately investigated and resolved, including with documentation in the literature if appropriate.

## Support

This study was funded by the National Institute for Health Research (NIHR) (Health and Social Care Delivery Research, Grant Reference Number NIHR 128364). The NIHR had no input in in study design, data collection, analysis, reporting, or the decision to submit for publication. For the purpose of Open Access, the author has applied a CC BY public copyright licence to any Author Accepted Manuscript version arising from this submission.

## Financial Disclosure

SJD has received research funding and lecture fees from Baxter Healthcare and Fresenius Medical Care (both companies deliver dialysis treatments, including home dialysis). ML has received research funding from Baxter Healthcare and speaker honoraria from Baxter Healthcare and Fresenius Medical Care. SJD and ML are members of the Behring LLC POSIBIL6ESKD steering committee. JF has received research funding from Baxter Healthcare.

## Acknowledgements

The kidney patient-level data reported here were supplied by the UKRR of the UK Kidney Association. We thank kidney patients and renal centres for providing data to the UKRR and for their participation in the national centre survey. The authors also thank the members of the Inter-CEPt Patient Advisory Group (PAG) for their input to and engagement with the study.

## Disclaimer

The views expressed are those of the authors and not necessarily those of the NIHR or the Department of Health and Social Care. The interpretation and reporting of the registry data are the responsibility of the authors and in no way should be seen as an official policy or interpretation of the UKRR or the UK Kidney Association.

## Supplementary Material

### Data aggregation, transformation, and survey question selection from the national survey of renal centres in England

The survey was open to all staff involved in providing home dialysis services at each kidney centre in England, including centre managers, clinical leads/directors, PD and HHD consultants and nurses, and Advanced Kidney Care (AKC) clinic staff and could be completed by multiple members of staff within the centre.

Responses to survey questions from individual staff at each centre were combined into a single centre-level response for each question. Pre-determined, pragmatic rules for data aggregation were followed to derive centre-level responses. Where multiple individuals had responded to a dichotomous or multiple-choice question from the same centre, the modal response was taken (e.g. if three respondents indicated that their centre offered peer support and one respondent reported that peer support was not offered or selected the ‘not sure’ option, that centre would be recorded as offering peer support). Where multiple individuals had responded but there was no unique modal response, the answer given by the most relevant staff role was taken (e.g. nurse responses relating to clinical practice were prioritised over responses from centre managers). In the case of Likert-type scale questions, if all responses were the same, there was full agreement, and that response was used. If the responses were different but there was a modal answer this was taken. If there was no modal response, then the median was calculated, if this was not a whole number this was rounded up. Following on from creating a single response, each Likert scale question was dichotomised (Scores of 0, 1 or 2 were combined to no/disagree and scored of 3 & 4 were combined to yes/agree)

Each survey question was then mapped against each centre to calculate the number and proportion of centres with missing data for each question. Initially, questions that applied to only a single HT modality (either home HD or PD individually) were excluded. This generated a list of potential candidate survey questions to take forward into the subsequent analysis. The final set of questions for the statistical modelling phase was decided by consensus, drawing on the literature, methodological and clinical expertise of team members to ensure that clinically relevant survey questions encompassing potentially modifiable elements of centre practices that could affect HT uptake were included, followed by exclusion of questions that had more than 10% missing data. An inspection of the pairwise correlation between the selected factors, revealed that “whether centres offer advice for special treatment registration” and “advice for working age patients” were perfectly correlated, therefore, only one of these was included in the analysis.

Figure S1 provides a flow diagram showing how the factors were selected from the survey.

**Figure S1:**
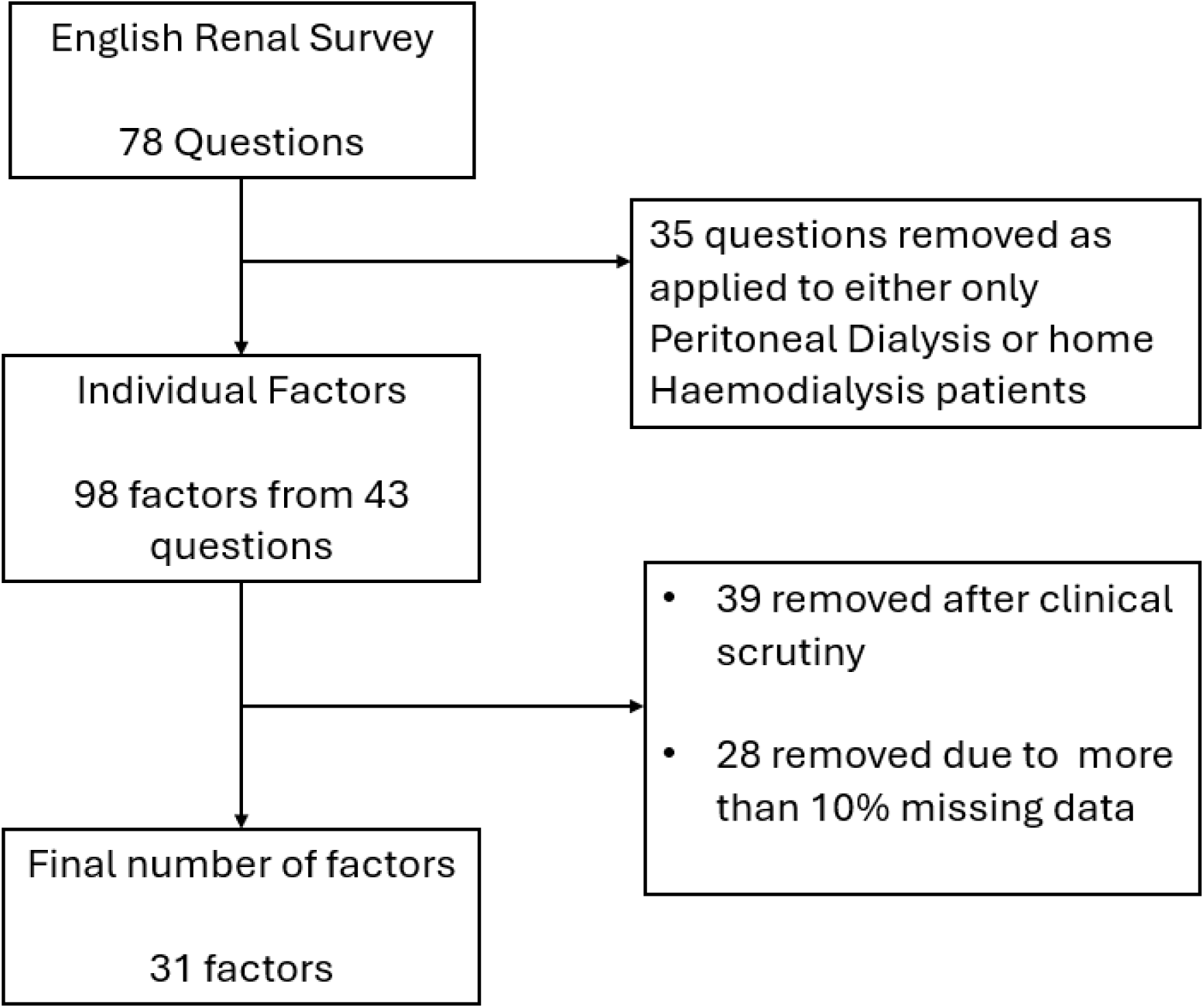
Flow diagram showing the selection of variables from the renal survey for inclusion in the sequences of regression analysis

### Supplementary information on sequences of regressions modelling

Sequences of regressions (SoR), a subclass of graphical models, is a multivariate statistical model that extend path analysis. They offer a novel approach to describing complex interrelations by simultaneously modelling multiple sets of outcome variables and demographic factors.

To examine the direct and indirect association between centre- and patient-level factors and the probability of HT uptake, the graphical model was specified by classifying the variables into sets of primary (HT uptake), intermediate (centre-level factors and patient characteristics) and demographic factors (Figure 2 of manuscript). The variables located on the left are response variables to those located to their right, and variables collocated within boxes are symmetrically associated. The model was built by fitting ordered sequences of logistic regression models, for each variable in the different groups as detailed in the Statistical Analysis section of the manuscript. The SoR model is characterised by a regression graph with nodes representing variables arranged in sets of primary, intermediate and demographic factors, connected by lines or arrow lines:

1. An arrow emerging from an explanatory variable and pointing to a response variable represents a directed association
2. A sequence of connected arrow lines linking two variables represents an indirect association, i.e., an association between two variables with intermediate explanatory variables
3. A dashed line connecting two variables collocated within the same group of intermediate variables represents a residual undirected association after controlling for the combined set of explanatory variables
4. A full line connecting two demographic variables represents an undirected association after controlling for other demographic variables

We note, however, that as the primary focus of the analysis was examining direct and indirect associations, we have not represented any undirected associations in the regression graphs. Therefore, an absence of a line linking two variables should not be interpreted as a lack of association. In fact, we explored undirected associations between the centre-level factors associated with HT uptake and found evidence of strong pairwise associations. However, these associations were not represented in the graphs to maintain clarity and avoid overcrowding (Figure 2 of manuscript).

Patterns of missingness were assessed. There were very low proportions of missing values across all the variables used in the SoR, ranging from 0% to 8% (median 0%, interquartile range 0% to 4%). This is unlikely to bias the parameters estimation which was based on the complete dataset.

While the model fitting required multiple statistical tests, the components of the model reflect distinctive relations of interest, arising from the hypothesised direction of associations between centre- and patient-level factors with HT uptake (Figure 1 of manuscript). Therefore, the interpretation of each statistical test performed is valid ^1^, and adjustment for multiple comparisons is not required. However, we used a level of statistical significance of 0.05 for model selection which is relatively low for a real-world data analysis.

All models were fitted using maximum likelihood estimation. As the model relies on local regression analyses, this allowed the identification of non-linear relations and checks of model assumptions. Diagnostic plots, including plots of residuals were used to check the model assumptions. The goodness of fit was assessed through the deviance statistic.

### Distribution of incident KRT patients from UKRR by centre and patterns of response by centre

Table S1 shows the number of patients who started KRT in each of the treatment centres by year. It also shows the difference in centre contribution to the number of patients in the dataset. Table S2 shows the demographics of the patients starting KRT and details the average number of patients starting treatment each year and the total number of patients who received HT within one year of starting KRT. Table S3 shows the patterns of response to each of the factors identified for inclusion in the SoR analysis.

**Table S1:**
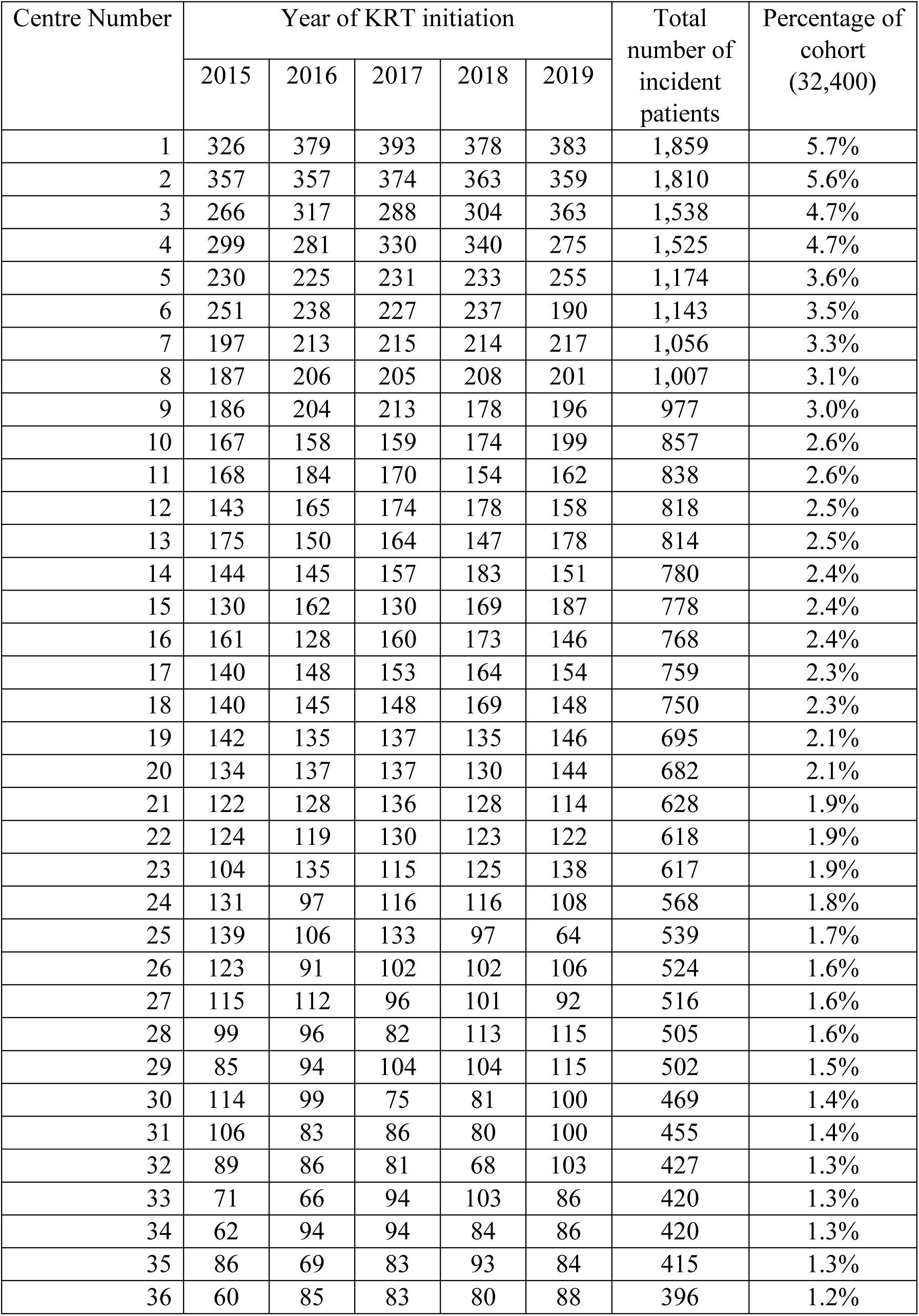
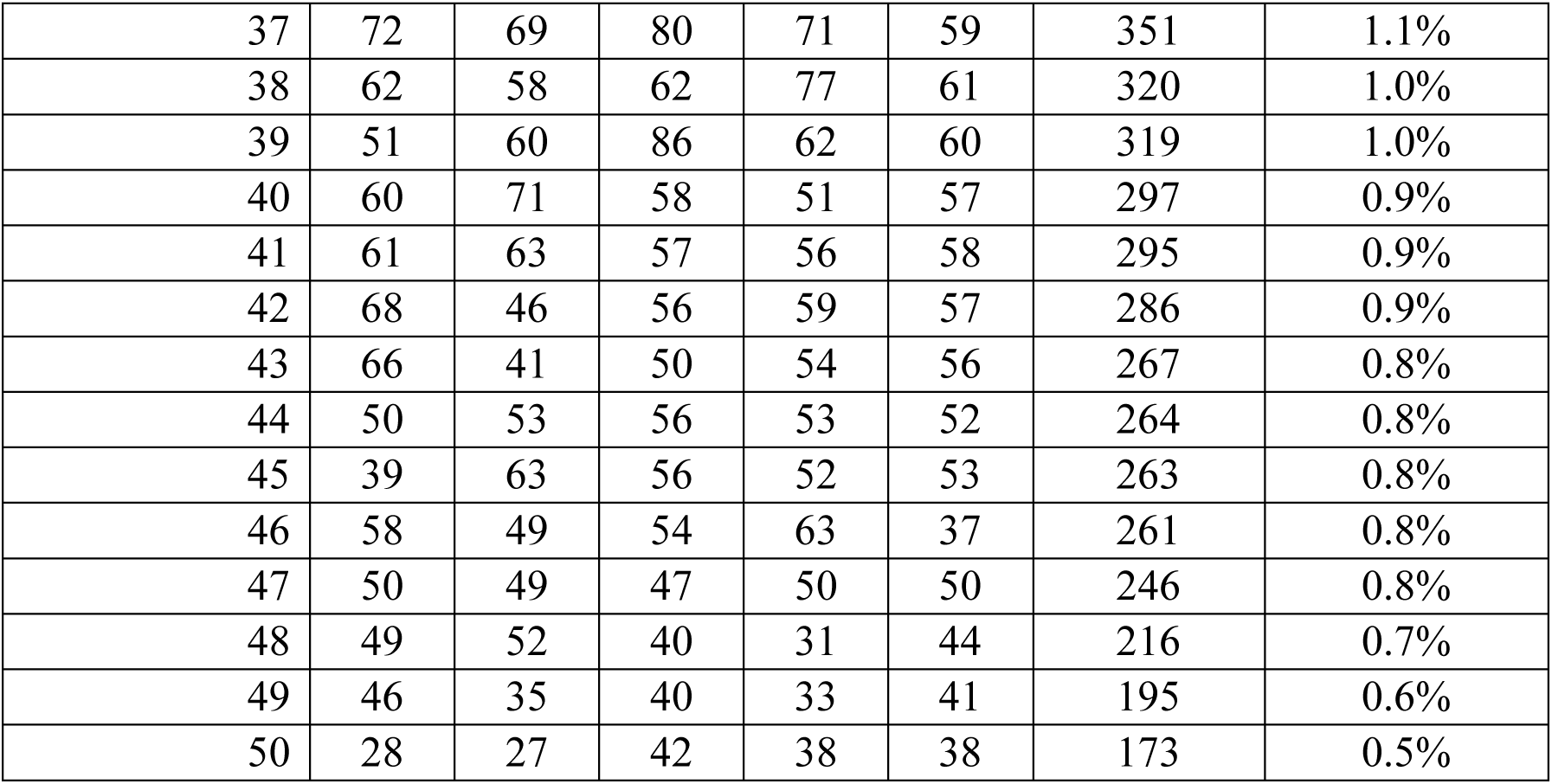
Number of incident patients starting KRT in each English renal centre from 2015-2019.

| Centre Number | Year of KRT initiation |  |  |  |  | Total number of incident patients | Percentage of cohort (32,400) |
| --- | --- | --- | --- | --- | --- | --- | --- |
|  | 2015 | 2016 | 2017 | 2018 | 2019 |  |  |
| 1 | 326 | 379 | 393 | 378 | 383 | 1,859 | 5.7% |
| 2 | 357 | 357 | 374 | 363 | 359 | 1,810 | 5.6% |
| 3 | 266 | 317 | 288 | 304 | 363 | 1,538 | 4.7% |
| 4 | 299 | 281 | 330 | 340 | 275 | 1,525 | 4.7% |
| 5 | 230 | 225 | 231 | 233 | 255 | 1,174 | 3.6% |
| 6 | 251 | 238 | 227 | 237 | 190 | 1,143 | 3.5% |
| 7 | 197 | 213 | 215 | 214 | 217 | 1,056 | 3.3% |
| 8 | 187 | 206 | 205 | 208 | 201 | 1,007 | 3.1% |
| 9 | 186 | 204 | 213 | 178 | 196 | 977 | 3.0% |
| 10 | 167 | 158 | 159 | 174 | 199 | 857 | 2.6% |
| 11 | 168 | 184 | 170 | 154 | 162 | 838 | 2.6% |
| 12 | 143 | 165 | 174 | 178 | 158 | 818 | 2.5% |
| 13 | 175 | 150 | 164 | 147 | 178 | 814 | 2.5% |
| 14 | 144 | 145 | 157 | 183 | 151 | 780 | 2.4% |
| 15 | 130 | 162 | 130 | 169 | 187 | 778 | 2.4% |
| 16 | 161 | 128 | 160 | 173 | 146 | 768 | 2.4% |
| 17 | 140 | 148 | 153 | 164 | 154 | 759 | 2.3% |
| 18 | 140 | 145 | 148 | 169 | 148 | 750 | 2.3% |
| 19 | 142 | 135 | 137 | 135 | 146 | 695 | 2.1% |
| 20 | 134 | 137 | 137 | 130 | 144 | 682 | 2.1% |
| 21 | 122 | 128 | 136 | 128 | 114 | 628 | 1.9% |
| 22 | 124 | 119 | 130 | 123 | 122 | 618 | 1.9% |
| 23 | 104 | 135 | 115 | 125 | 138 | 617 | 1.9% |
| 24 | 131 | 97 | 116 | 116 | 108 | 568 | 1.8% |
| 25 | 139 | 106 | 133 | 97 | 64 | 539 | 1.7% |
| 26 | 123 | 91 | 102 | 102 | 106 | 524 | 1.6% |
| 27 | 115 | 112 | 96 | 101 | 92 | 516 | 1.6% |
| 28 | 99 | 96 | 82 | 113 | 115 | 505 | 1.6% |
| 29 | 85 | 94 | 104 | 104 | 115 | 502 | 1.5% |
| 30 | 114 | 99 | 75 | 81 | 100 | 469 | 1.4% |
| 31 | 106 | 83 | 86 | 80 | 100 | 455 | 1.4% |
| 32 | 89 | 86 | 81 | 68 | 103 | 427 | 1.3% |
| 33 | 71 | 66 | 94 | 103 | 86 | 420 | 1.3% |
| 34 | 62 | 94 | 94 | 84 | 86 | 420 | 1.3% |
| 35 | 86 | 69 | 83 | 93 | 84 | 415 | 1.3% |
| 36 | 60 | 85 | 83 | 80 | 88 | 396 | 1.2% |
| 37 | 72 | 69 | 80 | 71 | 59 | 351 | 1.1% |
| 38 | 62 | 58 | 62 | 77 | 61 | 320 | 1.0% |
| 39 | 51 | 60 | 86 | 62 | 60 | 319 | 1.0% |
| 40 | 60 | 71 | 58 | 51 | 57 | 297 | 0.9% |
| 41 | 61 | 63 | 57 | 56 | 58 | 295 | 0.9% |
| 42 | 68 | 46 | 56 | 59 | 57 | 286 | 0.9% |
| 43 | 66 | 41 | 50 | 54 | 56 | 267 | 0.8% |
| 44 | 50 | 53 | 56 | 53 | 52 | 264 | 0.8% |
| 45 | 39 | 63 | 56 | 52 | 53 | 263 | 0.8% |
| 46 | 58 | 49 | 54 | 63 | 37 | 261 | 0.8% |
| 47 | 50 | 49 | 47 | 50 | 50 | 246 | 0.8% |
| 48 | 49 | 52 | 40 | 31 | 44 | 216 | 0.7% |
| 49 | 46 | 35 | 40 | 33 | 41 | 195 | 0.6% |
| 50 | 28 | 27 | 42 | 38 | 38 | 173 | 0.5% |

**Table S2:** Demographics of incident KRT patients in each English Renal Centre, from 2015-2019.

| Centre Number | No. of incident KRT patients | Average number of patients | Proportion of patients on HT within 1 year of KRT start | Sex | Ethnicity |  |  |  |  |  | Area of deprivation quintile |  |  |  |  |  | Patient waitlisted at start of KRT |  |  | Diabetes as primary diagnosis |  |  |
| --- | --- | --- | --- | --- | --- | --- | --- | --- | --- | --- | --- | --- | --- | --- | --- | --- | --- | --- | --- | --- | --- | --- |
|  |  |  |  | Males | Asian | Black | Mixed | Other | White | Missing | 1 (Least deprived) | 2 | 3 | 4 | 5 (Most deprived) | Missing | No | Yes | Pre-emptive transplant | No | Yes | Missing |
| 1 | 1,859 | 372 | 16% | 63% | 40% | 18% | 0% | 1% | 41% | 0% | 10% | 15% | 26% | 33% | 16% | 0% | 82% | 10% | 8% | 55% | 45% | 0% |
| 2 | 1,810 | 362 | 31% | 61% | 27% | 10% | 2% | 1% | 57% | 3% | 9% | 10% | 16% | 16% | 48% | 0% | 84% | 10% | 6% | 69% | 29% | 3% |
| 3 | 1,538 | 308 | 21% | 64% | 17% | 5% | 1% | 1% | 69% | 7% | 18% | 20% | 19% | 23% | 20% | 0% | 78% | 13% | 9% | 58% | 22% | 20% |
| 4 | 1,525 | 305 | 36% | 63% | 32% | 20% | 13% | 4% | 29% | 2% | 4% | 8% | 18% | 42% | 29% | 0% | 84% | 10% | 6% | 48% | 33% | 19% |
| 5 | 1,174 | 235 | 32% | 64% | 21% | 19% | 8% | 0% | 42% | 9% | 7% | 16% | 22% | 31% | 24% | 0% | 76% | 15% | 9% | 60% | 31% | 9% |
| 6 | 1,143 | 229 | 18% | 63% | 15% | 10% | 2% | 2% | 64% | 7% | 32% | 19% | 20% | 21% | 7% | 0% | 86% | 9% | 5% | 83% | 12% | 5% |
| 7 | 1,056 | 211 | 29% | 64% | 3% | 1% | 1% | 1% | 80% | 14% | 23% | 22% | 24% | 18% | 12% | 0% | 76% | 14% | 9% | 43% | 16% | 41% |
| 8 | 1,007 | 201 | 24% | 63% | 8% | 3% | 1% | 2% | 66% | 19% | 29% | 27% | 21% | 13% | 9% | 0% | 69% | 14% | 17% | 64% | 27% | 9% |
| 9 | 977 | 195 | 26% | 62% | 14% | 19% | 2% | 0% | 61% | 3% | 12% | 12% | 12% | 20% | 43% | 0% | 79% | 10% | 11% | 56% | 26% | 18% |
| 10 | 857 | 171 | 14% | 60% | 9% | 28% | 2% | 2% | 53% | 6% | 14% | 12% | 20% | 32% | 21% | 1% | 77% | 12% | 12% | 55% | 26% | 19% |
| 11 | 838 | 168 | 35% | 63% | 16% | 3% | 1% | 1% | 79% | 0% | 10% | 12% | 13% | 18% | 46% | 0% | 73% | 14% | 13% | 68% | 30% | 2% |
| 12 | 818 | 164 | 21% | 63% | 17% | 5% | 0% | 2% | 76% | 0% | 12% | 15% | 14% | 19% | 40% | 0% | 72% | 16% | 12% | 71% | 28% | 0% |
| 13 | 814 | 163 | 32% | 64% | 14% | 32% | 1% | 5% | 46% | 2% | 10% | 15% | 20% | 33% | 22% | 0% | 88% | 9% | 3% | 65% | 35% | 0% |
| 14 | 780 | 156 | 18% | 67% | 7% | 3% | 0% | 2% | 84% | 4% | 11% | 15% | 17% | 17% | 41% | 0% | 84% | 11% | 5% | 73% | 27% | 1% |
| 15 | 778 | 156 | 17% | 67% | 11% | 5% | 3% | 2% | 65% | 13% | 21% | 20% | 23% | 25% | 11% | 0% | 82% | 12% | 6% | 72% | 27% | 1% |
| 16 | 768 | 154 | 21% | 61% | 15% | 1% | 0% | 0% | 84% | 0% | 12% | 19% | 13% | 21% | 35% | 0% | 75% | 13% | 12% | 72% | 27% | 0% |
| 17 | 759 | 152 | 21% | 65% | 4% | 6% | 0% | 2% | 86% | 3% | 21% | 21% | 19% | 19% | 19% | 0% | 79% | 13% | 8% | 71% | 28% | 2% |
| 18 | 750 | 150 | 23% | 64% | 5% | 1% | 2% | 1% | 85% | 7% | 19% | 24% | 26% | 16% | 15% | 0% | 85% | 8% | 7% | 81% | 19% | 0% |
| 19 | 695 | 139 | 19% | 63% | 4% | 1% | 1% | 0% | 91% | 3% | 13% | 21% | 21% | 24% | 22% | 0% | 85% | 8% | 7% | 77% | 22% | 1% |
| 20 | 682 | 136 | 27% | 66% | 1% | 1% | 0% | 3% | 94% | 1% | 13% | 18% | 32% | 27% | 10% | 0% | 86% | 7% | 7% | 80% | 20% | 0% |
| 21 | 628 | 126 | 24% | 64% | 4% | 1% | 0% | 1% | 93% | 0% | 14% | 13% | 16% | 25% | 32% | 0% | 78% | 10% | 12% | 78% | 22% | 0% |
| 22 | 618 | 124 | 31% | 64% | 11% | 6% | 1% | 1% | 81% | 0% | 16% | 14% | 17% | 19% | 34% | 0% | 78% | 11% | 12% | 77% | 23% | 0% |
| 23 | 617 | 123 | 29% | 62% | 16% | 4% | 0% | 0% | 79% | 0% | 16% | 25% | 20% | 21% | 18% | 0% | 81% | 11% | 8% | 69% | 23% | 7% |
| 24 | 568 | 114 | 19% | 68% | 6% | 1% | 0% | 0% | 93% | 0% | 13% | 15% | 14% | 17% | 40% | 0% | 81% | 10% | 9% | 72% | 27% | 1% |
| 25 | 539 | 108 | 23% | 63% | 4% | 4% | 1% | 2% | 86% | 5% | 11% | 11% | 12% | 18% | 47% | 2% | 75% | 9% | 16% | 63% | 23% | 14% |
| 26 | 524 | 105 | 30% | 66% | 3% | 1% | 0% | 1% | 95% | 0% | 14% | 18% | 15% | 22% | 31% | 0% | 82% | 11% | 7% | 78% | 22% | 0% |
| 27 | 516 | 103 | 32% | 63% | 5% | 1% | 1% | 1% | 89% | 4% | 18% | 22% | 16% | 21% | 23% | 0% | 85% | 11% | 4% | 64% | 30% | 6% |
| 28 | 505 | 101 | 15% | 65% | 4% | 1% | 1% | 1% | 85% | 9% | 24% | 23% | 31% | 15% | 5% | 2% | 69% | 9% | 22% | 84% | 14% | 2% |
| 29 | 502 | 100 | 31% | 64% | 19% | 5% | 0% | 2% | 60% | 14% | 32% | 17% | 21% | 25% | 5% | 0% | 80% | 10% | 10% | 68% | 31% | 1% |
| 30 | 469 | 94 | 22% | 68% | 1% | 0% | 1% | 0% | 96% | 1% | 13% | 20% | 24% | 21% | 21% | 0% | 89% | 6% | 5% | 73% | 21% | 6% |
| 31 | 455 | 91 | 22% | 63% | 23% | 21% | 1% | 12% | 35% | 8% | 24% | 21% | 27% | 21% | 8% | 0% | 83% | 9% | 8% | 60% | 29% | 11% |
| 32 | 427 | 85 | 17% | 62% | 41% | 2% | 0% | 2% | 54% | 1% | 4% | 10% | 12% | 21% | 53% | 0% | 84% | 10% | 6% | 65% | 34% | 1% |
| 33 | 420 | 84 | 21% | 68% | 2% | 0% | 1% | 1% | 93% | 2% | 19% | 28% | 25% | 18% | 10% | 0% | 82% | 12% | 6% | 70% | 25% | 4% |
| 34 | 420 | 84 | 22% | 59% | 4% | 0% | 0% | 0% | 94% | 0% | 9% | 15% | 12% | 26% | 38% | 0% | 88% | 7% | 5% | 71% | 29% | 0% |
| 35 | 415 | 83 | 29% | 65% | 23% | 9% | 1% | 1% | 65% | 0% | 7% | 14% | 14% | 19% | 46% | 0% | 91% | 8% | 1% | 68% | 32% | 0% |
| 36 | 396 | 79 | 53% | 64% | 10% | 3% | 1% | 2% | 84% | 1% | 15% | 17% | 23% | 15% | 29% | 0% | 82% | 14% | 4% | 67% | 32% | 1% |
| 37 | 351 | 70 | 30% | 67% | 3% | 2% | 1% | 1% | 93% | 0% | 30% | 19% | 28% | 14% | 10% | 0% | 85% | 11% | 5% | 71% | 28% | 1% |
| 38 | 320 | 64 | 37% | 63% | 4% | 1% | 2% | 0% | 92% | 2% | 15% | 25% | 24% | 25% | 11% | 0% | 90% | 7% | 3% | 77% | 23% | 0% |
| 39 | 319 | 64 | 30% | 68% | 1% | 0% | 1% | 2% | 96% | 0% | 8% | 16% | 23% | 33% | 20% | 0% | 82% | 10% | 8% | 75% | 24% | 1% |
| 40 | 297 | 59 | 28% | 66% | 1% | 0% | 0% | 1% | 95% | 3% | 32% | 25% | 25% | 10% | 9% | 0% | 80% | 10% | 10% | 77% | 23% | 0% |
| 41 | 295 | 59 | 21% | 58% | 2% | 0% | 0% | 1% | 95% | 1% | 14% | 22% | 13% | 21% | 32% | 0% | 91% | 8% | 2% | 66% | 32% | 2% |
| 42 | 286 | 57 | 18% | 62% | 0% | 0% | 1% | 0% | 99% | 0% | 3% | 14% | 30% | 38% | 15% | 0% | 84% | 11% | 5% | 71% | 29% | 0% |
| 43 | 267 | 53 | 31% | 64% | 2% | 2% | 1% | 13% | 74% | 8% | 17% | 24% | 30% | 13% | 16% | 0% | 85% | 8% | 7% | 53% | 10% | 37% |
| 44 | 264 | 53 | 30% | 64% | 12% | 5% | 1% | 0% | 83% | 0% | 8% | 17% | 12% | 23% | 40% | 0% | 88% | 8% | 4% | 76% | 24% | 0% |
| 45 | 263 | 53 | 22% | 62% | 4% | 1% | 0% | 1% | 94% | 0% | 6% | 14% | 20% | 25% | 34% | 0% | 87% | 10% | 2% | 70% | 29% | 1% |
| 46 | 261 | 52 | 28% | 61% | 0% | 0% | 0% | 0% | 98% | 2% | 9% | 12% | 16% | 15% | 48% | 0% | 87% | 11% | 2% | 74% | 26% | 0% |
| 47 | 246 | 49 | 27% | 69% | 5% | 5% | 4% | 1% | 81% | 3% | 22% | 14% | 13% | 29% | 21% | 0% | 91% | 6% | 2% | 66% | 32% | 2% |
| 48 | 216 | 43 | 30% | 71% | 5% | 2% | 0% | 1% | 85% | 6% | 25% | 29% | 25% | 18% | 3% | 0% | 83% | 13% | 5% | 69% | 30% | 1% |
| 49 | 195 | 39 | 38% | 62% | 0% | 0% | 0% | 0% | 100% | 0% | 10% | 15% | 24% | 32% | 20% | 0% | 88% | 7% | 5% | 71% | 26% | 3% |
| 50 | 173 | 35 | 1% | 64% | 1% | 1% | 0% | 2% | 94% | 2% | 10% | 18% | 20% | 22% | 29% | 0% | 94% | 6% | 0% | 63% | 31% | 6% |

**Table S3:**
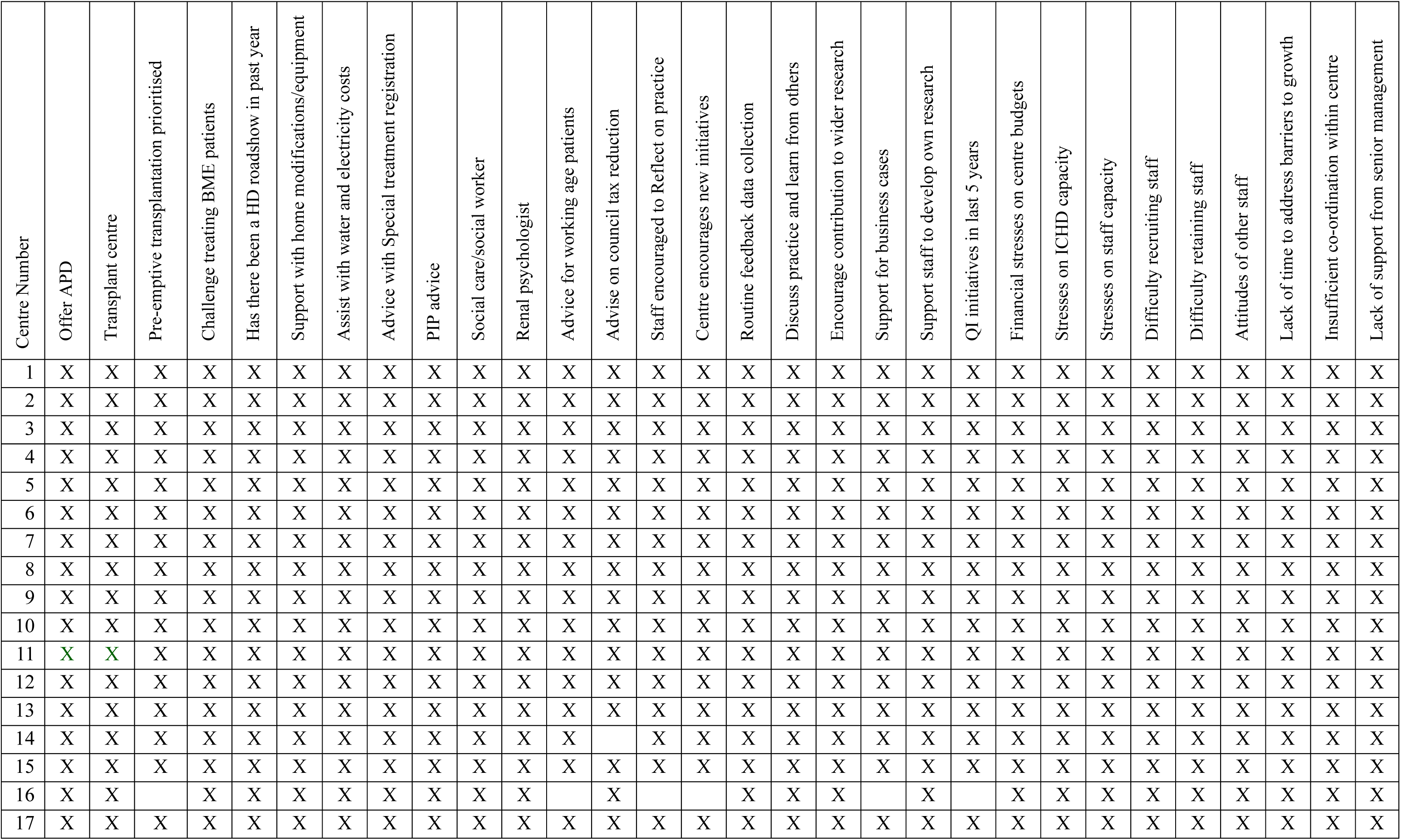
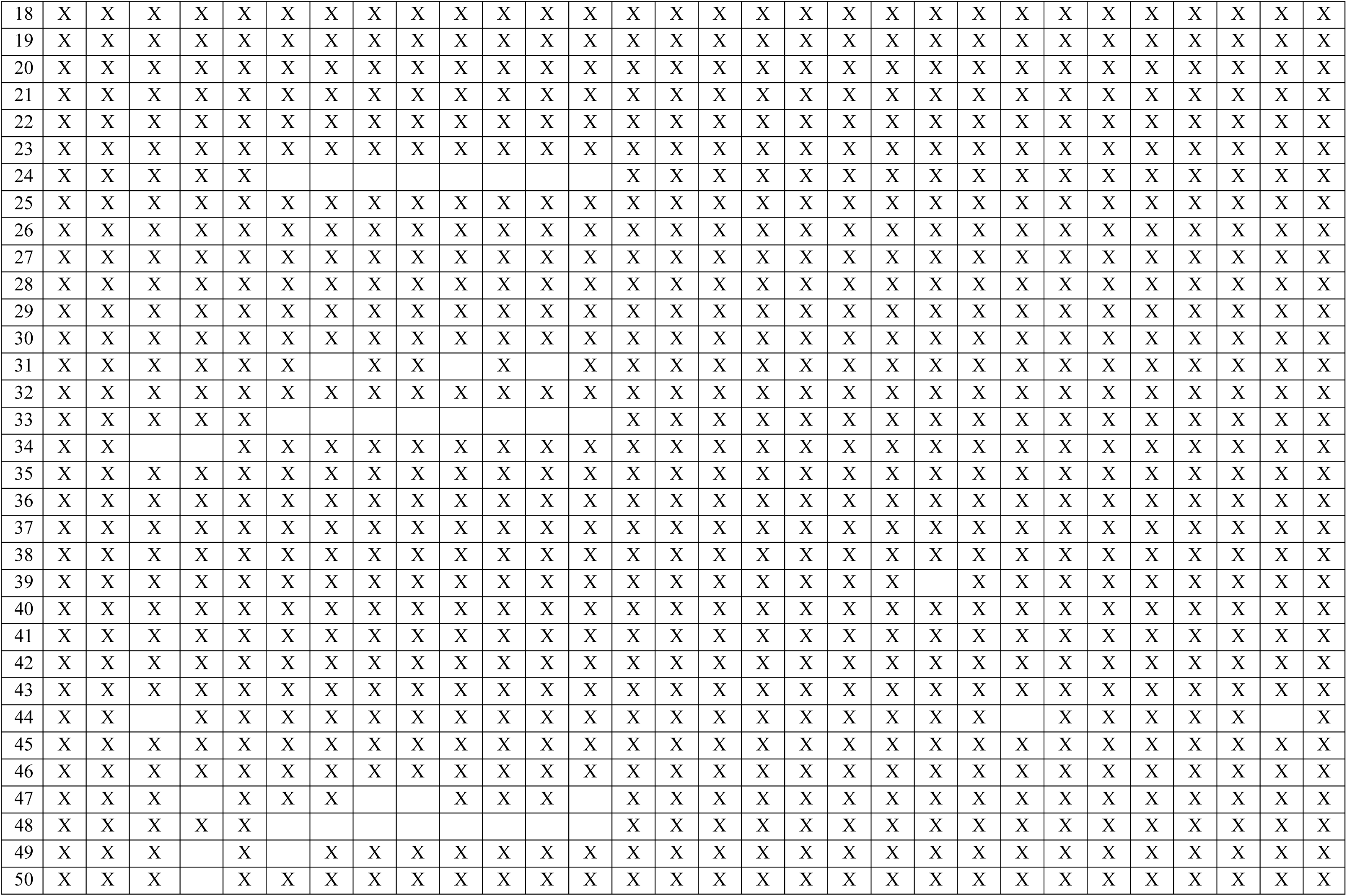
Patterns of centre response to each of the centre level variables selected for inclusion in the sequence of regressions analysis.

### Predicted probabilities of HT update for centre- and patient-level factors that were identified as being directly associated with HT uptake

For each of the centre level (Figure S2) and patient level (Figures S3 and S4) factors that were identified as being directly associated with HT uptake, the predicted probability of uptake along with 95% confidence interval was calculated for each of the effects in turn.

**Figure S2:**
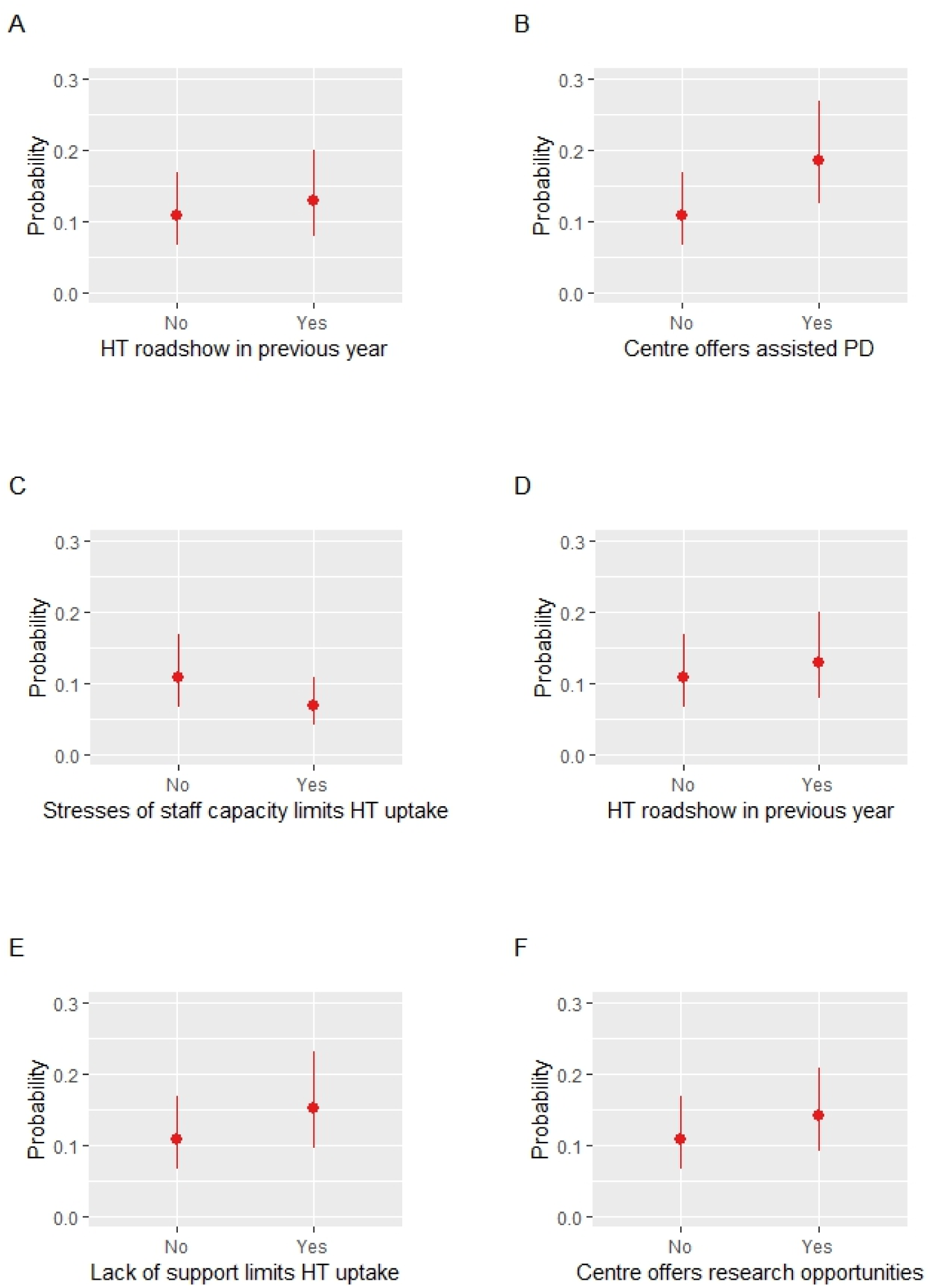
Predicted probability of HT uptake for each centre-level variables that were identified to influence HT uptake directly

**Figure S3:**
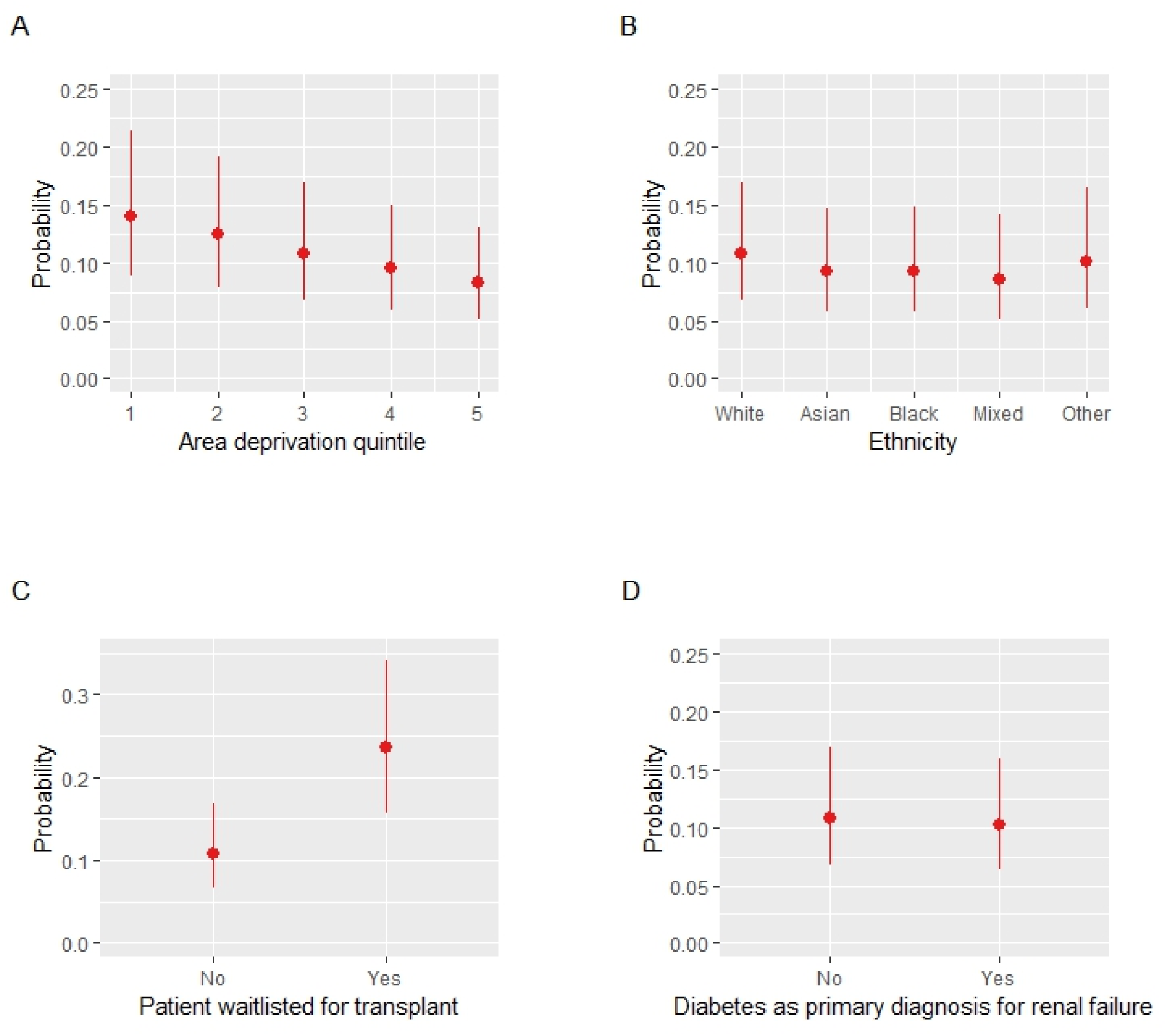
Predicted probability of HT uptake for categorical patient demographics/characteristics that influence HT uptake directly

**Figure S4:**
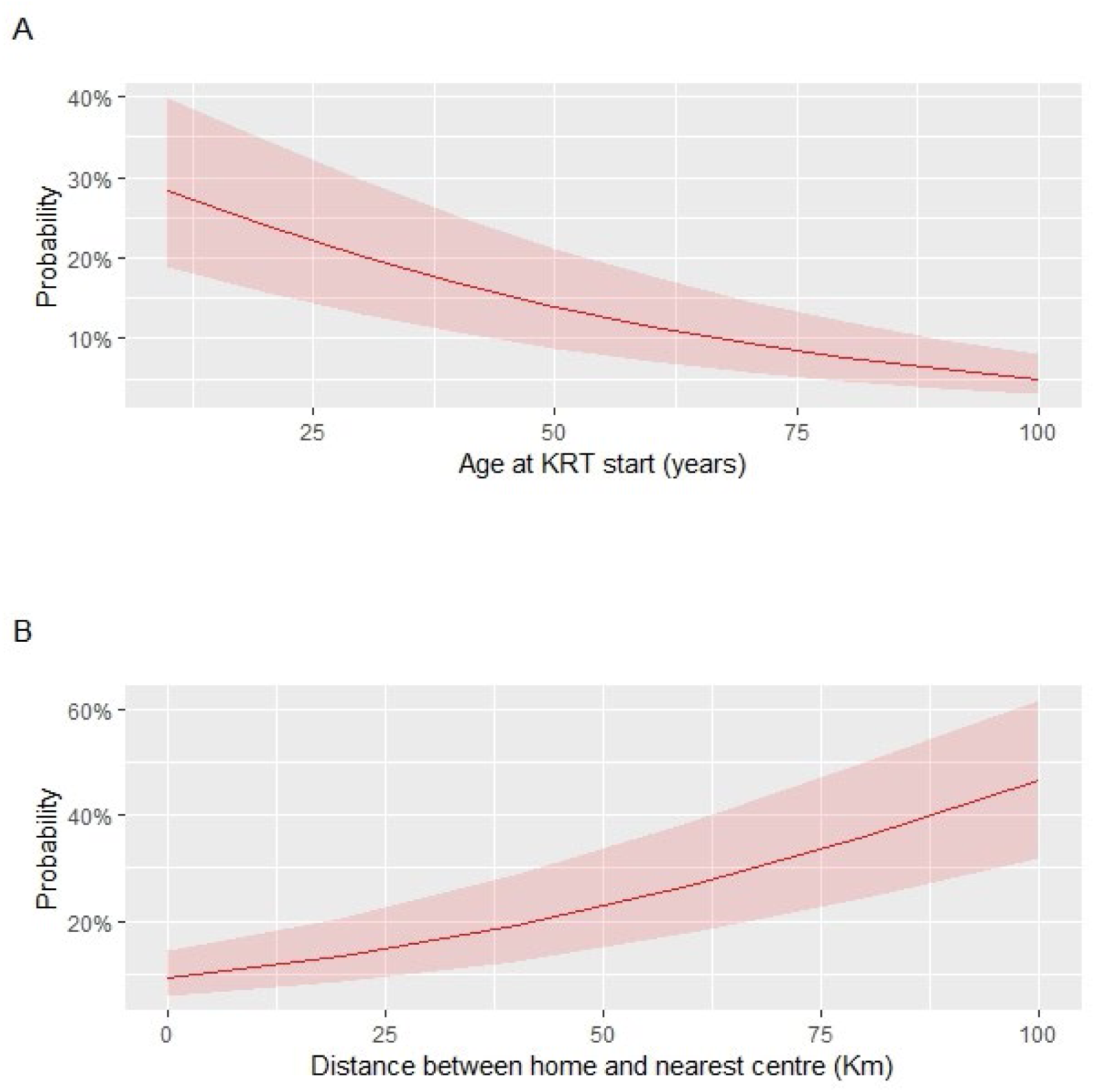
Predicted probability of HT uptake for continuous patient demographics that influence HT uptake directly

### Interaction terms on the mixed-effects logistic regression model for HT uptake

We identified two potential interaction terms: between research opportunities and perceived lack of support limiting HT and between research opportunities and running QI projects. Due to data sparsity, inclusion of these interactions leads to wide intervals for the main effects (See Table S4). The wide confidence intervals indicate a high degree of uncertainty, and the findings should be interpreted with caution. Figure S5 suggests that the predicted probability of HT uptake may be higher for patients in centres which provide research opportunities, however this effect may be amplified if there is perceived lack of support which limits HT. If a patient is attached to a renal centre that does not provide research opportunities for staff but has had QI initiatives within the past 5 years, the probability of HT uptake may be higher than for a patient in a centre where there have been no QI initiatives. When a centre provides staff with research opportunities the difference in the probability of HT uptake decreases but remains higher for centres with QI initiatives (Figure S6)

**Table S4:** Odds Ratios and 95% CI for model including potential interaction terms between research opportunities and QI initiatives and research opportunities and lack of support on HT uptake.

|  | OR (95% CI) |
| --- | --- |
| Centre encourages opportunities for wider research | 28.54 (3.40-239.83) |
| QI initiatives with past 5 years | 46.31 (5.73-374.23) |
| Lack of Support limits HT access | 1.01 (0.67-1.51) |
| Centre encourages opportunities for wider research x QI initiatives with past 5 years | 0.03 (0.01-0.28) |
| Centre encourages opportunities for wider research x Lack of Support limits HT access | 1.73 (1.09-2.74) |

**Figure S5:**
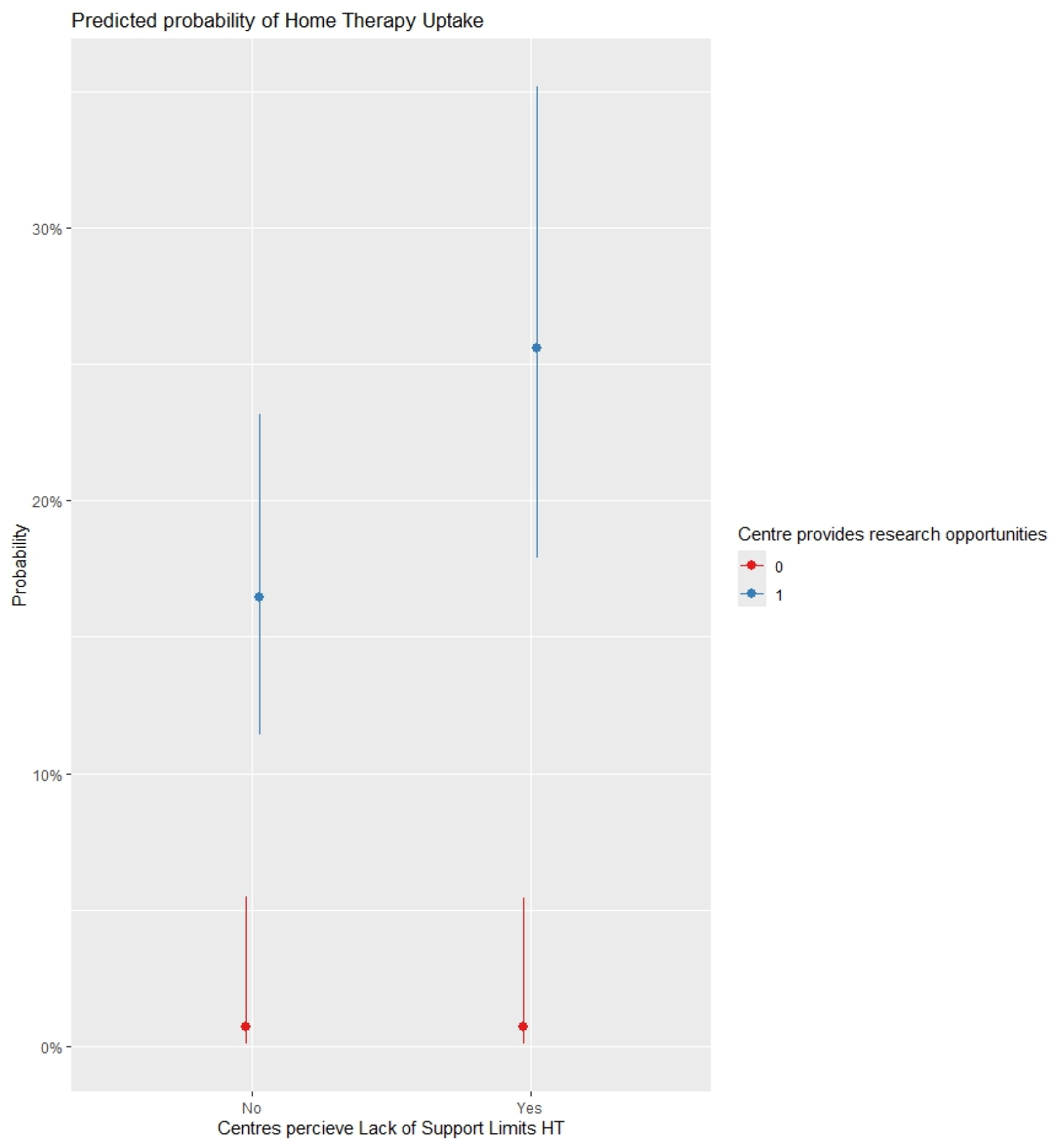
Predicted probability (95% CI) of HT uptake for the interaction between lack of support limiting access to home therapy and centres providing staff with opportunities for research

**Figure S6:**
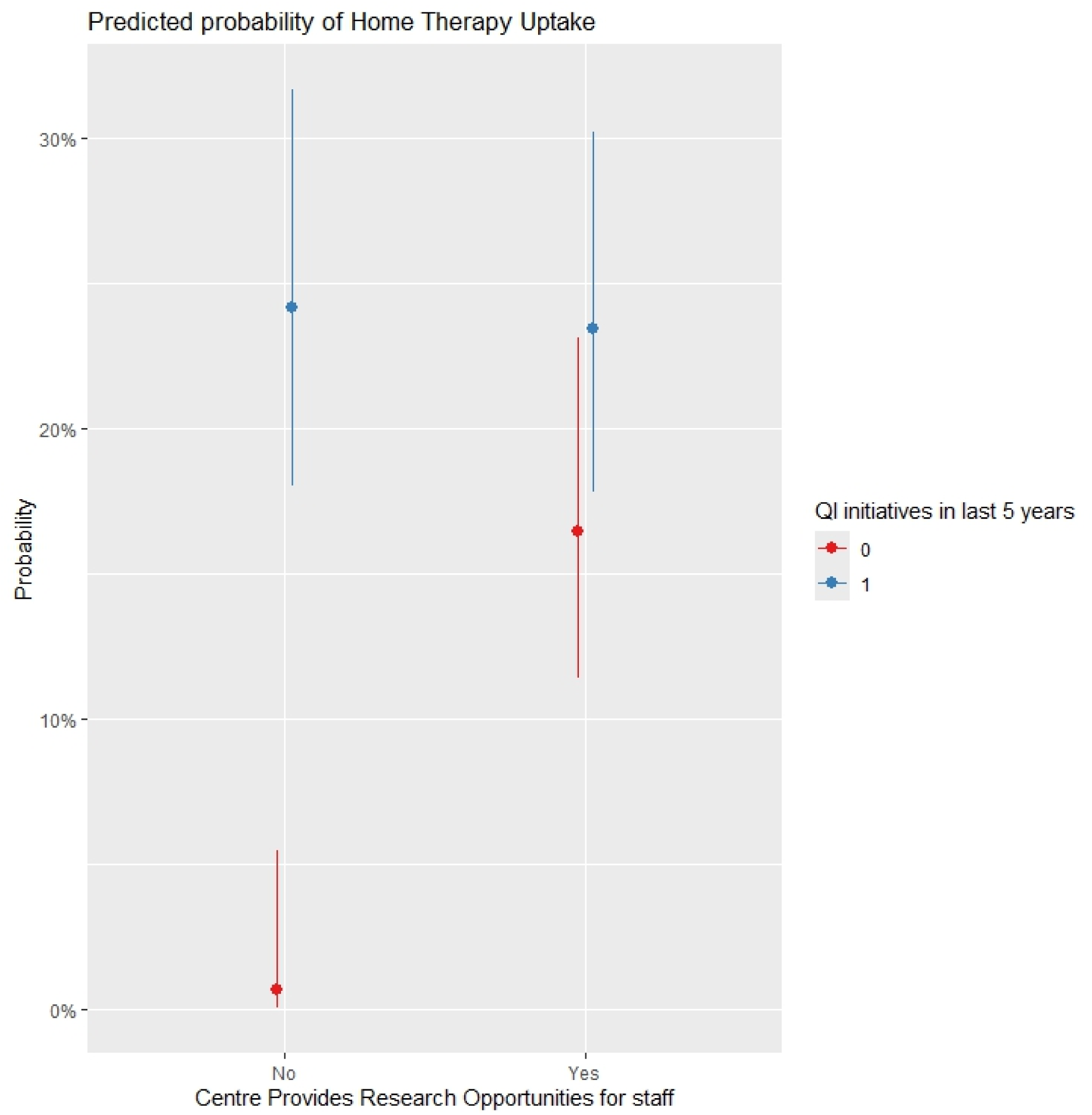
Predicted probability (95% CI) of HT uptake for the interaction between QI initiatives with past 5 years and centres providing staff with research opportunities

### Regression models for centre-level and patient-level factors

For ease of presentation, Figure S7 shows subgraphs displaying the associations between patient characteristics and centre-level factors as response variables, grouping them within stacked boxes if they had the same explanatory factors. This does not mean that the effect of the explanatory factors was the same. Tables S4 presents the full results from the regression models for centre-level factors and Table S5 presents the regression models fitted were patient-level factors were response variables. For all centre levels factors, there are differences between different ethnic groups and patients from different levels of neighbourhood deprivation. There is only one centre level factor that is influenced by patient sex. For centres where there are difficulties recruiting staff, which limits access to HT there are fewer females than males OR [95%CI] (0.93 [0.88-0.99]) Table S4.

**Figure S7:**
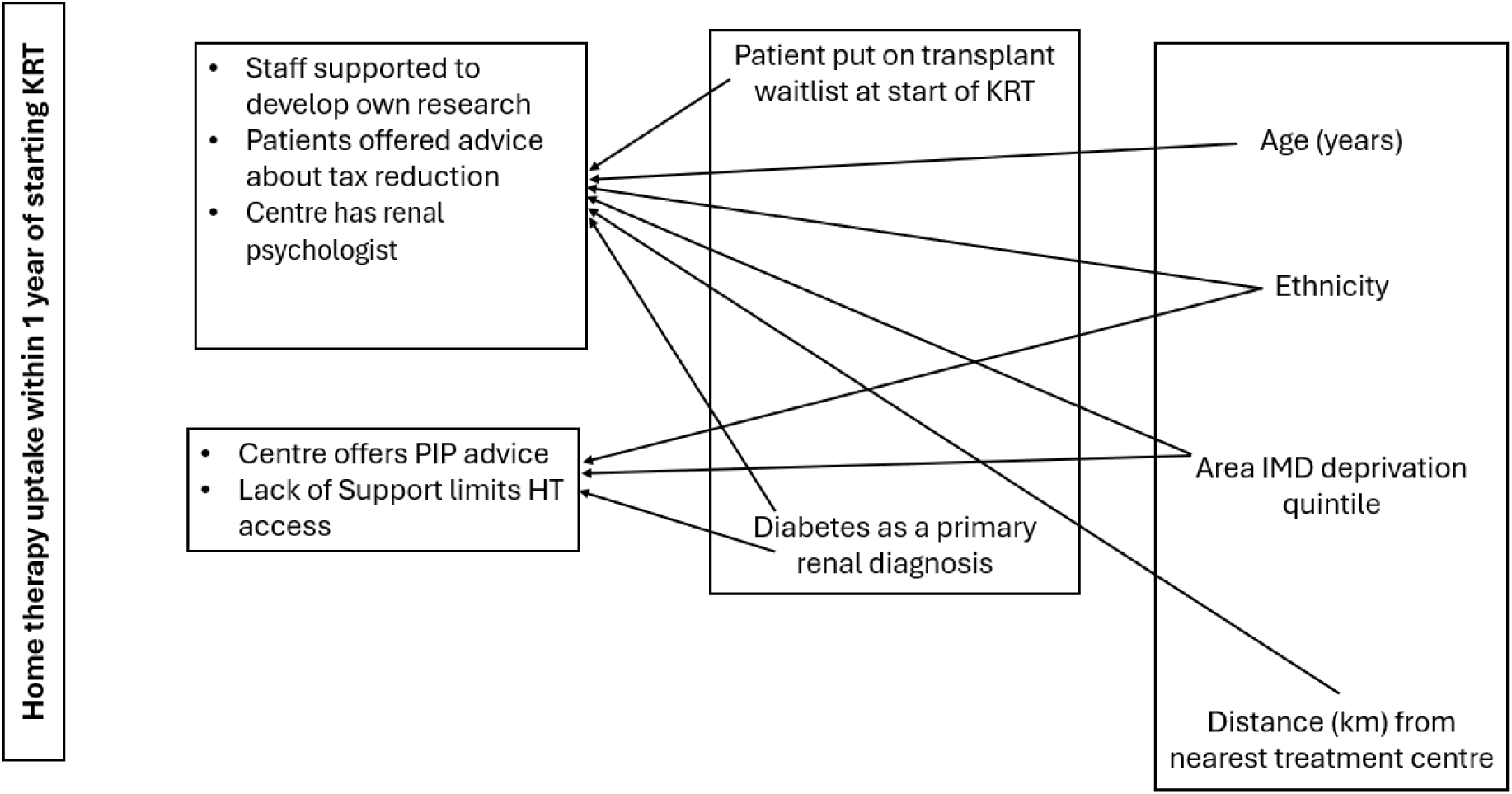
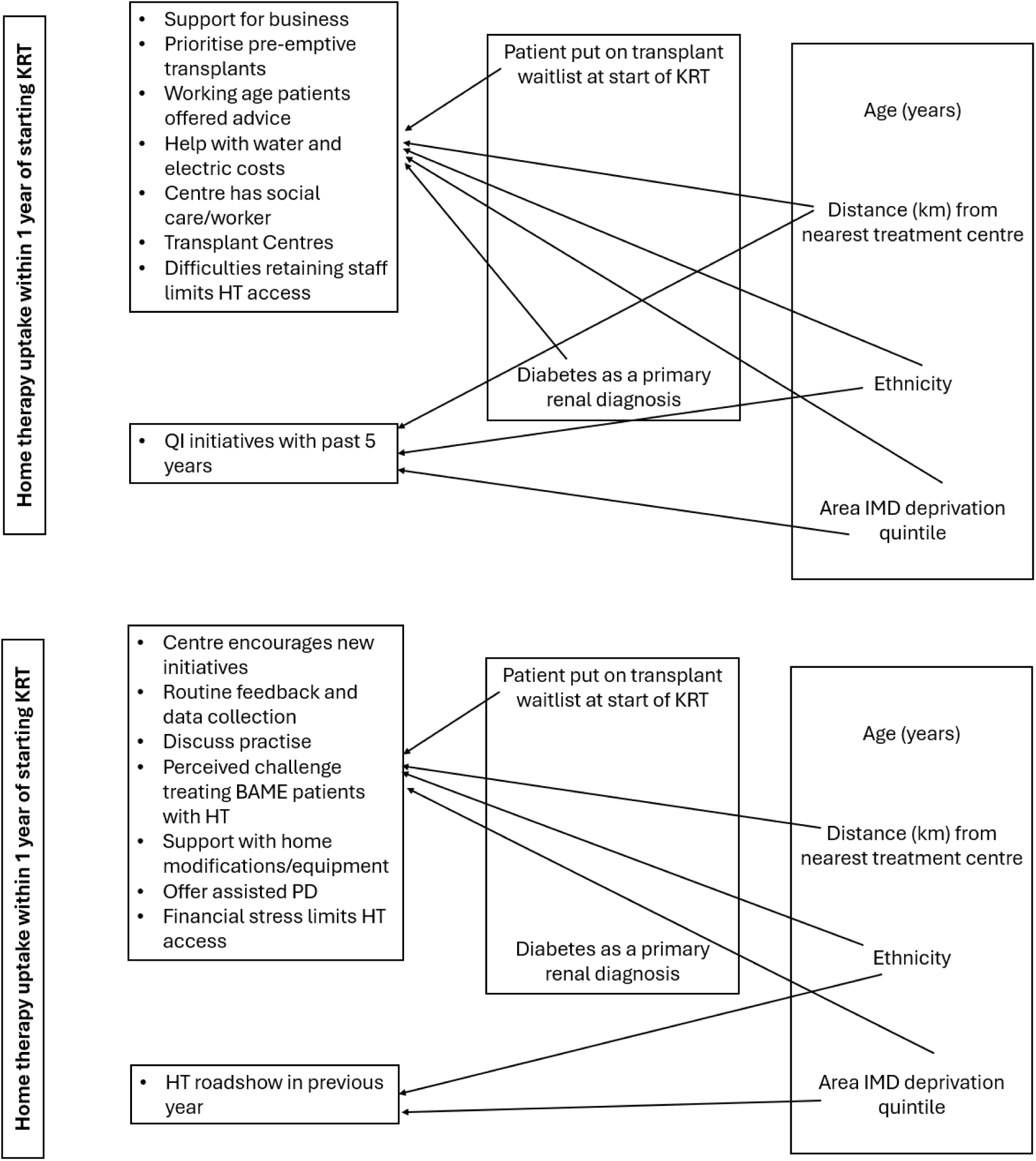
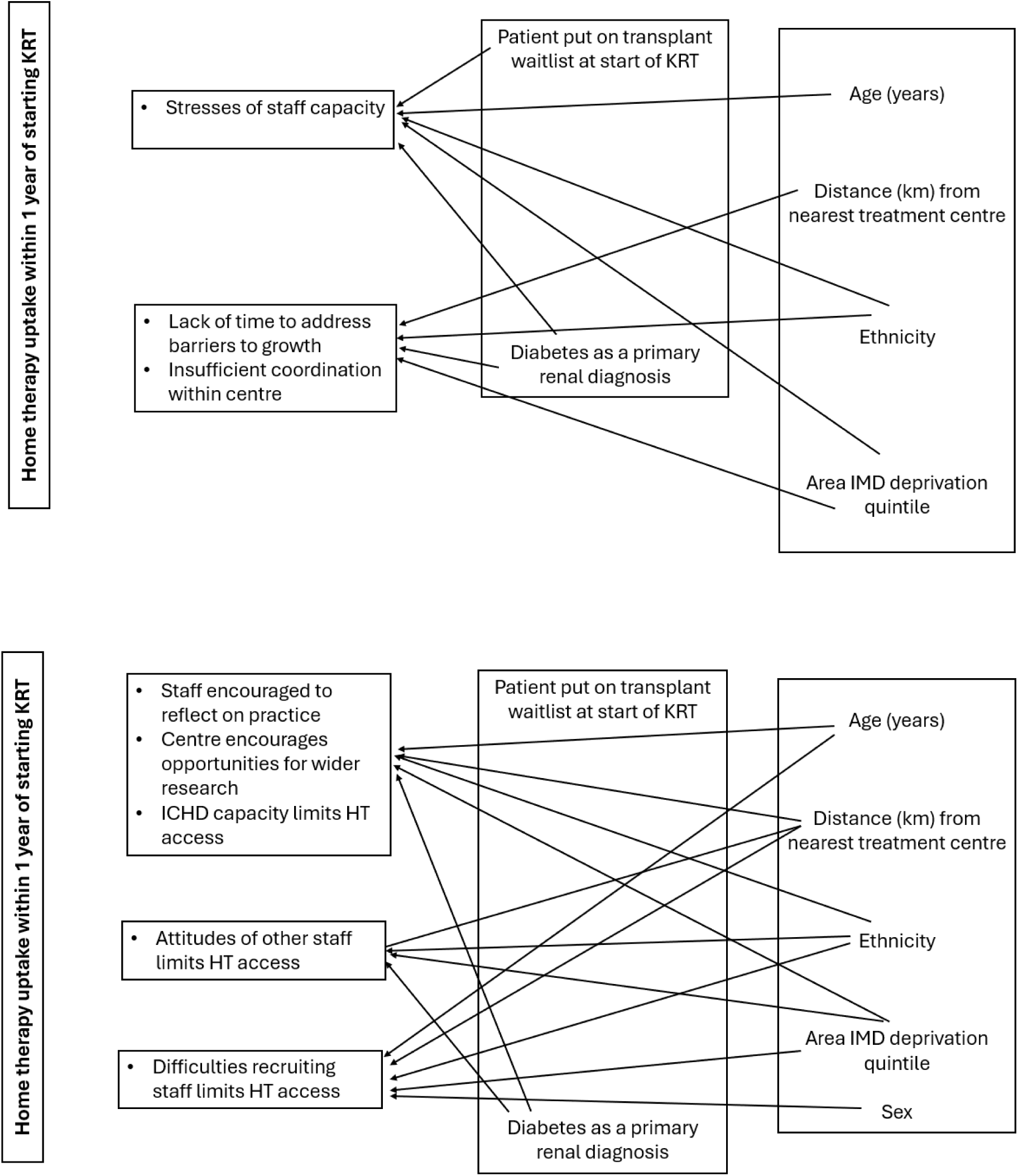
Regression subgraph for direct associations between patient characteristics and centre-level factors. The direction and strength of associations are shown by odds ratios (95% CI) provided in Table S4.

**Table S5:** Regression models for centre level factors as response variables (Figure S7) based on the logistic regression models of best fit, Odds Ratios (95% CI).

| Centre level variable | Explanatory variables |  |  |  |  |  |  |
| --- | --- | --- | --- | --- | --- | --- | --- |
|  | Patient waitlisted for transplant at start of KRT | Diabetes as primary renal diagnosis | Distance (per 10km) from nearest centre | Age (per 10-year increase at start of KRT) | Ethnicity | IMD deprivation level quintile<br>1 – Least deprived<br>5 – Most deprived | Sex |
| Staff supported to develop own research | 1.09 (1.01-1.18) | 1.11 (1.05-1.17) | 0.92 (0.91-0.94) | 1.01 (1.00-1.02) | Asian - 1.40 (1.29-1.51)<br>Black - 1.26 (1.14-1.38)<br>Mixed - 0.64 (0.53-0.78)<br>Other - 0.64 (0.53-0.77) | 1 - 0.82 (0.75-0.89)<br>2 - 1.06 (0.98-1.15)<br>4 - 0.90 (0.84-0.97)<br>5 - 1.09 (1.02-1.18) |  |
| Patients offered advice about tax reduction | 1.22 (1.05-1.42) | 0.65 (0.59-0.71) | 1.18 (1.14-1.22) | 0.98 (0.97-1.00) | Asian - 0.38 (0.34-0.43)<br>Black - 0.49 (0.42-0.56)<br>Mixed - 8.50 (3.17-22.84)<br>Other - 1.37 (0.90-2.08) | 1 - 1.63 (1.38-1.94)<br>2 - 1.21 (1.04-1.40)<br>4 - 1.05 (0.93-1.19)<br>5 - 1.92 (1.68-2.20) |  |
| Centre has renal psychologist | 1.17 (1.05-1.29) | 0.74 (0.69-0.79) | 0.96 (0.94-0.97) | 0.97 (0.96-0.98) | Asian - 0.86 (0.79-0.94)<br>Black - 1.17 (1.04-1.33)<br>Mixed - 3.20 (2.21-4.63)<br>Other - 1.31 (1.01-1.69) | 1 - 1.22 (1.10-1.36)<br>2 - 1.12 (1.01-1.23)<br>4 - 0.94 (0.86-1.02)<br>5 - 1.61 (1.46-1.77) |  |
| Support for business cases | 1.15 (1.05-1.27) | 1.32 (1.24-1.42) | 0.76 (0.75-0.77) |  | Asian - 1.40 (1.26-1.54)<br>Black - 1.83 (1.60-2.11)<br>Mixed - 1.40 (1.07-1.83)<br>Other - 1.21 (0.95-1.55) | 1 - 0.88 (0.80-0.97)<br>2 - 1.17 (1.06-1.29)<br>4 - 1.02 (0.93-1.12)<br>5 - 1.58 (1.44-1.74) |  |
| Prioritise pre-emptive transplants | 1.17 (1.04-1.33) | 1.42 (1.30-1.56) | 0.95 (0.93-0.98) |  | Asian - 1.06 (0.94-1.20)<br>Black - 1.37 (1.16-1.62)<br>Mixed - 1.65 (1.13-2.42)<br>Other - 1.59 (1.12-2.28) | 1 - 0.70 (0.62-0.80)<br>2 - 0.85 (0.76-0.94)<br>4 - 1.13 (1.00-1.27)<br>5 - 1.39 (1.23-1.57) |  |
| Working age patients offered advice | 1.73 (1.30-2.29) | 0.85 (0.73-0.99) | 0.88 (0.84-0.91) |  | Asian - 4.46 (3.10-6.40)<br>Black - 5.50 (3.33-9.08)<br>Mixed - 2.06 (1.02-4.17)<br>Other - 2.32 (1.09-4.93) | 1 - 0.87 (0.66-1.15)<br>2 - 0.81 (0.62-1.05)<br>4 - 0.56 (0.44-0.71)<br>5 - 0.44 (0.35-0.56) |  |
| Help with water and electric costs | 1.51 (1.17-1.95) | 0.83 (0.71-0.97) | 0.88 (0.85-0.92) |  | Asian - 0.94 (0.75-1.18)<br>Black - 1.39 (0.99-1.96) | 1 - 0.50 (0.40-0.63)<br>2 - 0.86 (0.68-1.10) |  |
|  |  |  |  |  | Mixed - 1.21 (0.62-2.37)<br>Other - 0.83 (0.47-1.46) | 4 - 0.80 (0.64-1.01)<br>5 - 1.38 (1.08-1.78) |  |
| Centre has social care/worker | 0.89 (0.81-0.96) | 0.90 (0.84-0.95) | 0.93 (0.92-0.95) |  | Asian - 1.41 (1.29-1.53)<br>Black - 1.70 (0.51-1.90)<br>Mixed - 0.55 (0.50-0.67)<br>Other - 0.86 (0.69-1.05) | 1 - 1.04 (0.95-1.15)<br>2 - 1.06 (0.97-1.17)<br>4 - 0.83 (0.76-0.90)<br>5 - 0.96 (0.88-1.04) |  |
| Transplant Centres | 1.35 (1.25-1.45) | 1.10 (1.04-1.16) | 0.88 (0.87-0.90) |  | Asian - 1.59 (1.48-1.71)<br>Black - 1.92 (1.75-2.10)<br>Mixed - 0.88 (0.73-1.07)<br>Other - 1.00 (0.83-1.20) | 1 - 0.98 (0.89-1.06)<br>2 - 0.98 (0.90-1.06)<br>4 - 0.89 (0.82-0.96)<br>5 - 0.96 (0.90-1.04) |  |
| Difficulties retaining staff limits HT access | 0.91 (0.84-0.99) | 0.81 (0.77-0.96) | 0.97 (0.96-0.99) |  | Asian - 1.15 (1.07-1.25)<br>Black - 2.17 (1.94-2.43)<br>Mixed - 6.77 (4.69-9.77)<br>Other - 2.60 (2.03-3.34) | 1 - 1.20 (1.09-1.32).<br>2 - 0.96 (0.88-1.04)<br>4 - 0.86 (0.79-0.93)<br>5 - 0.76 (0.70-0.82) |  |
| Stresses on staff capacity limits HT access | 1.16 (1.05-1.28) | 1.18 (1.10-1.26) |  | 0.99 (0.98-0.99) | Asian - 1.15 (1.05-1.26)<br>Black - 2.25 (1.97-2.59)<br>Mixed - 1.72(1.30-2.28)<br>Other - 1.84 (1.40-2.43) | 1 - 1.34 (1.19-1.51)<br>2 - 1.06 (0.95-1.18)<br>4 - 0.93 (0.84-1.02)<br>5 - 0.51 (0.47-0.56) |  |
| Centre encourages new initiatives | 0.87 (0.77-0.98) |  | 0.77 (0.75-0.78) |  | Asian - 1.16 (1.01-1.33)<br>Black - 2.09 (1.68-2.60)<br>Mixed - 1.80 (1.19-2.73)<br>Other - 1.69 (1.15-2.50) | 1 - 0.84 (0.74-0.96)<br>2 - 1.03 (0.91-1.78)<br>4 - 1.04 (0.92-1.18)<br>5 - 1.00 (0.88-1.14) |  |
| Routine feedback and data collection | 1.11 (1.02-1.21) |  | 0.89 (0.87-0.90) |  | Asian - 1.27 (1.17-1.39)<br>Black - 0.85 (0.77-0.94)<br>Mixed - 0.40 (0.33-0.48)<br>Other - 0.81 (0.67-0.99) | 1 - 1.32 (1.20-1.45)<br>2 - 1.21 (0.11-1.32)<br>4 - 0.90 (0.83-0.98)<br>5 - 1.47 (0.35-1.60) |  |
| Discuss practise | 1.18 (1.06-1.30) |  | 0.92 (0.90-0.94) |  | Asian - 0.76 (0.69-0.84)<br>Black - 0.46 (0.42-0.52)<br>Mixed - 0.22 (0.18-0.27)<br>Other - 0.55 (0.44-0.68) | 1 - 1.39 (1.23 -1.56)<br>2 - 1.29 (1.16-1.44)<br>4 - 0.83 (0.76-0.91)<br>5 - 1.62 (1.47-1.80) |  |
| Perceived challenge treating BAME patients with HT | 0.86 (0.77-0.95) |  | 0.95 (0.93-0.97) |  | Asian - 2.17 (2.00-2.35)<br>Black - 1.50 (1.40-1.73)<br>Mixed - 4.36 (1.60-5.26)<br>Other - 1.99 (1.62-2.45) | 1 - 0.76 (0.67-0.86)<br>2 - 0.86 (0.77-0.97)<br>4 - 1.22 (1.11-1.35)<br>5 - 1.66 (1.51-1.83) |  |
| Support with home modifications/equipment | 1.13 (1.02-1.25) |  | 0.94 (0.92-0.96) |  | Asian - 0.90 (0.82-1.00)<br>Black - 0.80 (0.71-0.90)<br>Mixed - 0.26 (0.22-0.32)<br>Other - 0.27 (0.22-0.33) | 1 - 1.30 (1.16-1.46)<br>2 - 1.18 (1.06-1.31)<br>4 - 0.90 (0.82-0.99)<br>5 - 1.49 (1.34-1.64) |  |
| Offer assisted PD | 1.15 (1.00-1.31) |  | 0.78 (0.77-0.79) |  | Asian - 4.12 (3.35-5.09)<br>Black - 12.05 (7.73-18.79)<br>Mixed - 3.49 (2.08-5.86)<br>Other - 3.37 (2.07-5.49) | 1 - 1.16 (1.09-1.44)<br>2 - 1.03 (0.91-1.17)<br>4 - 0.99 (0.87-1.12)<br>5 - 0.80 (0.78-1.00) |  |
| Financial stress limits HT access | 0.84 (0.78-0.91) |  | 0.96 (0.95-0.97) |  | Asian - 1.05 (0.98-1.13)<br>Black - 1.43 (1.31-1.57)<br>Mixed - 0.39 (0.31-0.50)<br>Other - 0.84 (0.70-1.02) | 1 - 1.21 (1.11-1.32)<br>2 - 1.04 (0.96-1.13)<br>4 - 0.95 (0.88-1.02)<br>5 - 0.77 (0.71-0.83) |  |
| Lack of time to address barriers to growth |  | 1.08 (1.02-1.14) | 0.89 (0.88-0.91) |  | Asian - 1.20 (1.11-1.30)<br>Black - 2.07 (1.87-2.30)<br>Mixed - 2.47 (1.97-3.09)<br>Other - 1.93 (1.57-2.39) | 1 - 1.01 (0.93-1.10)<br>2 - 1.00 (0.92-1.08)<br>4 - 0.95 (0.88-1.03)<br>5 - 0.56 (0.52-0.61) |  |
| Staff encouraged to reflect on practice |  | 0.87 (0.78-0.96) | 0.95 (0.93-0.98) | 1.03 (1.02-1.04) | Asian - 1.49 (1.26-1.77)<br>Black - 0.48 (0.41-0.86)<br>Mixed - 0.64 (0.47-0.88)<br>Other - 0.64 (0.47-0.88) | 1 - 0.78 (0.67-0.91)<br>2 - 0.98 (0.84-1.14)<br>4 - 0.97 (0.77-1.02)<br>5 - 1.70 (1.46-1.99) |  |
| Centre encourages opportunities for wider research |  | 0.93 (0.86-0.99) | 0.96 (0.94-0.97) | 0.99 (0.98-1.00) | Asian - 3.73 (3.25-4.28)<br>Black - 1.59 (1.39-1.81)<br>Mixed - 2.20 (1.61-3.02)<br>Other - 1.61 (1.23-2.13) | 1 - 1.07 (0.96-1.20)<br>2 - 1.02 (0.92-1.14)<br>4 - 0.86 (0.78-0.95)<br>5 - 0.75 (0.68-0.83) |  |
| ICHHD capacity limits HT access |  | 1.20 (1.14-1.26) | 0.96 (0.94-0.97) | 0.99 (0.98-0.99) | Asian - 1.42 (1.32-1.52)<br>Black - 1.67 (1.52-1.83)<br>Mixed - 2.88 (2.31-3.58)<br>Other - 1.11 (0.93-1.34) | 1 - 0.88 (0.82-0.96)<br>2 - 1.03 (0.95-1.11)<br>4 - 1.01 (0.94-1.07)<br>5 - 0.68 (0.63-0.73) |  |
| Insufficient coordination within centre |  | 1.07 (1.01-1.13) | 0.97 (0.97-0.98) |  | Asian - 1.86 (1.73-2.00)<br>Black - 1.84 (1.67-2.01)<br>Mixed - 1.91 (1.59-2.30)<br>Other - 1.45 (1.20-1.75) | 1 - 1.08 (1.00-1.18)<br>2 - 0.99 (0.91-1.07)<br>4 - 1.01 (0.93-1.08)<br>5 - 0.65 (0.60-0.70) |  |
| Centre offers PIP advice |  | 0.82 (0.73-0.92) |  |  | Asian - 5.74 (4.41-7.48)<br>Black - 7.76 (5.21-11.58) | 1 - 0.91 (0.74-1.11)<br>2 - 0.76 (0.63-0.92) |  |
|  |  |  |  |  | Mixed - 4.04 (2.08-7.83)<br>Other - 2.11 (1.29-3.43) | 4 - 0.70 (0.58-0.94)<br>5 - 0.44 (0.37-0.52) |  |
| Lack of Support limits HT access |  | 1.09 (1.02-1.16) |  |  | Asian - 0.98 (0.90-1.06)<br>Black - 1.64 (1.49-1.81)<br>Mixed - 5.14 (4.26-6.20)<br>Other - 0.90 (0.72-1.12) | 1 - 0.81 (0.74-0.89)<br>2 - 0.92 (0.84-1.00)<br>4 - 1.03 (0.95-1.11)<br>5 - 0.71-0.65-0.77) |  |
| Difficulties recruiting staff limits HT access |  |  | 0.94 (0.93-0.96) | 0.99 (0.98-0.99) | Asian - 1.24 (1.14-1.35)<br>Black - 3.05 (2.66-3.52)<br>Mixed (2.68 (2.02-3.57)<br>Other - 4.03- (2.87-5.70) | 1 - 1.30 (1.17-1.45)<br>2 - 0.98 (0.89-1.08)<br>4 - 0.84 (0.76-0.92)<br>5 - 0.42-0.69-0.46) | Female- 0.93 (0.88-0.99) |
| Attitudes of other staff limits HT access |  |  | 0.96 (0.94-0.97) | 0.99 (0.98-0.99) | Asian - 1.35 (1.26-1.44)<br>Black - 1.07 (0.99-1.17)<br>Mixed - 1.34 (1.13-1.59)<br>Other - 1.19 (0.99-1.41) | 1 - 1.24 (1.15-1.34)<br>2 - 1.04 (0.96-1.12)<br>4 - 0.96 (0.90-1.02)<br>5 - 0.76 (0.71-0.81) |  |
| QI initiatives with past 5 years |  |  | 0.88 (0.86-0.90) |  | Asian - 2.43 (2.03-2.91)<br>Black - 3.27 (2.50-4.27)<br>Mixed - 1.58 (1.07-2.31)<br>Other - 1.78 (0.90-1.81) | 1 - 1.00 (0.87-1.15)<br>2 - 0.96 (0.84-1.09)<br>4 - 1.02 (0.90-1.17)<br>5 - 1.40 (1.22-1.61) |  |
| HT roadshow* in previous year |  |  |  |  | Asian - 1.20 (1.05-1.37)<br>Black - 1.17 (0.98-1.39)<br>Mixed - 0.61 (0.38-0.97)<br>Other - 0.87 (0.58-1.29) | 1 - 0.95 (0.81-1.12)<br>2 - 0.91 (0.77-1.06)<br>4 - 1.07 (0.93-1.23)<br>5 - 0.76 (0.66-0.89) |  |

\*An HT roadshow is an initiative whereby a dialysis centre is visited by a team including patients, family members, clinicians and industry to promote HTs; IMD: Index of Multiple Deprivation; HT: Home therapy; KRT: Kidney replacement therapy; PD: Peritoneal dialysis; QI: Quality improvement; PIP; Personal Independence payment

**Table S6:** Regression models for patient-level factors as response variables based on the logistic regression models of best fit, Odds Ratios (95% CI).

|  | Explanatory Variable |  |  |  |
| --- | --- | --- | --- | --- |
| Patient characteristics | Age (per 10-year increase at start of KRT) | Ethnicity | IMD deprivation level quintile<br>1 – Least deprived<br>5 – Most deprived | Sex |
| Diabetes as primary renal diagnosis | 1.02 (1.01-1.03) | Asian - 2.52 (2.35-2.70)<br>Black - 1.54 (1.40-1.69)<br>Mixed - 1.49 (1.22-1.81)<br>Other - 1.24 (1.01-1.51) | 1 - 0.74 (0.67-0.82)<br>2 - 0.91 (0.83-0.99)<br>4 - 1.22 (1.12-1.32)<br>5 - 1.30 (1.19-1.40) | Female - 0.94 (0.88-0.98) |
| Patient waitlisted at start of KRT | 0.82 (0.81-0.83) | Asian - 1.42 (1.28-1.56)<br>Black - 0.96 (0.84-1.10)<br>Mixed - 1.09 (0.83-1.42)<br>Other - 0.74 (0.54-1.00) | 1 - 1.47 (1.30-1.67)<br>2 - 1.15 (1.02-1.30)<br>4 - 0.78 (0.70-0.88)<br>5 - 0.69 (0.62-0.77) |  |

